# Repurposing digitised clinical narratives to discover prognostic factors and predict survival in patients with advanced cancer

**DOI:** 10.1101/2020.10.28.20214627

**Authors:** Frank PY Lin, Osama SM Salih, Nina Scott, Michael B Jameson, Richard J Epstein

## Abstract

Electronic medical records (EMR) represent a rich informatics resource that remains largely unexploited for improving healthcare outcomes. Here we report a systematic text mining analysis of EMR correspondence for 4791 cancer patients treated between 2001 and 2017. Meaningful groups of text descriptors correlating with poor survival outcomes were systematically identified, and applying machine learning analysis to clinical text accurately predicted cancer patient survival at selected timepoints up to 12 months. In a validation cohort of 726 patients, inclusion of EMR descriptors to machine learning models outperformed the predictivity of conventional clinical symptom scores by 4.9% (*p* = 0.001). These results prove that labour-intensive EMR data collection can be repurposed to add clinical value. Extension of this approach to a broader spectrum of digital health data should transform the real-time utility of such latent informatics resources, enabling healthcare systems to be more adaptive and responsive to patient circumstances.

---

Predicting disease behaviour is a key decision-making variable in modern cancer management. To this end, quantifying the benefits of costly target-specific therapeutics has become more complex with the growth of biomarker technology [1,2]. Although certain clinical correlates, such as patient performance status, are well-validated [3], other factors often affect treatment outcomes – such as comorbidities [4], germline genetic variations [5] or resource constraints [6]. Candidate prognostic factors can only be assessed if prospectively included in study designs [7,8], while patient survival predictions based on clinician judgement often prove inaccurate [9-11]. Development of more sophisticated decision support tools is thus a crucial research priority for precision medicine [12].

To survey relevant variables in patients with advanced cancer, we used an *a posteriori* informatics strategy, leveraging unstructured text from routine electronic medical records (EMR). Lead-up work confirmed that valid predictive factors could be extracted by mining EMRs using Text-based Exploratory Pattern Analyser for Prognosticator and Associator discovery (TEPAPA) [13]. Prognostic variable identification was addressed by hypothesising that interpretable text patterns associated with survival are non-randomly documented by oncologists. Though many nuanced phrasings may not be recognised *a priori*, this contrasts with retrospective use of codified registry data limited to pre-defined variables. For the present study, we hypothesised that incorporation of survival analysis to guide descriptor discovery can yield useful algorithms (see Online Methods).

To explore the textual landscape relevant to advanced cancers, we screened 17067 patients across all cancer types presenting to a tertiary cancer centre in New Zealand between January 2001 and April 2017; over 7000 of these patients had advanced incurable cancer (Stage IV), 4791 of whom (68%) had at least one EMR document retrievable for analysis. The characteristics of this study cohort, listed in Table 1, include age, gender, survival, ethnicity, cancer type, and stage. Seventeen patients had metachronous cancers of different histology. Median follow-up was 55.6 months (95% CI: 50.7–59.8). Median overall survival (OS) was 13.5 months (95% CI: 12.8–14.2), being shortest in pancreatic (4.6 months, 2.8–6.4), unknown primary (6.3 months, 4.7–8.7), gastro-oesophageal (6.9 months, 5.6–8.5) and non-small-cell lung (7.4 months, 6.5–8.3) cancers. Head and neck squamous cell cancer (HNSCC) patients enjoyed the longest OS (63.7 months; 95% CI: 40.3– 133.0), though this result may have been confounded by inclusion of stage IV patients who received curative-intent treatment [14].

**Table 1.**
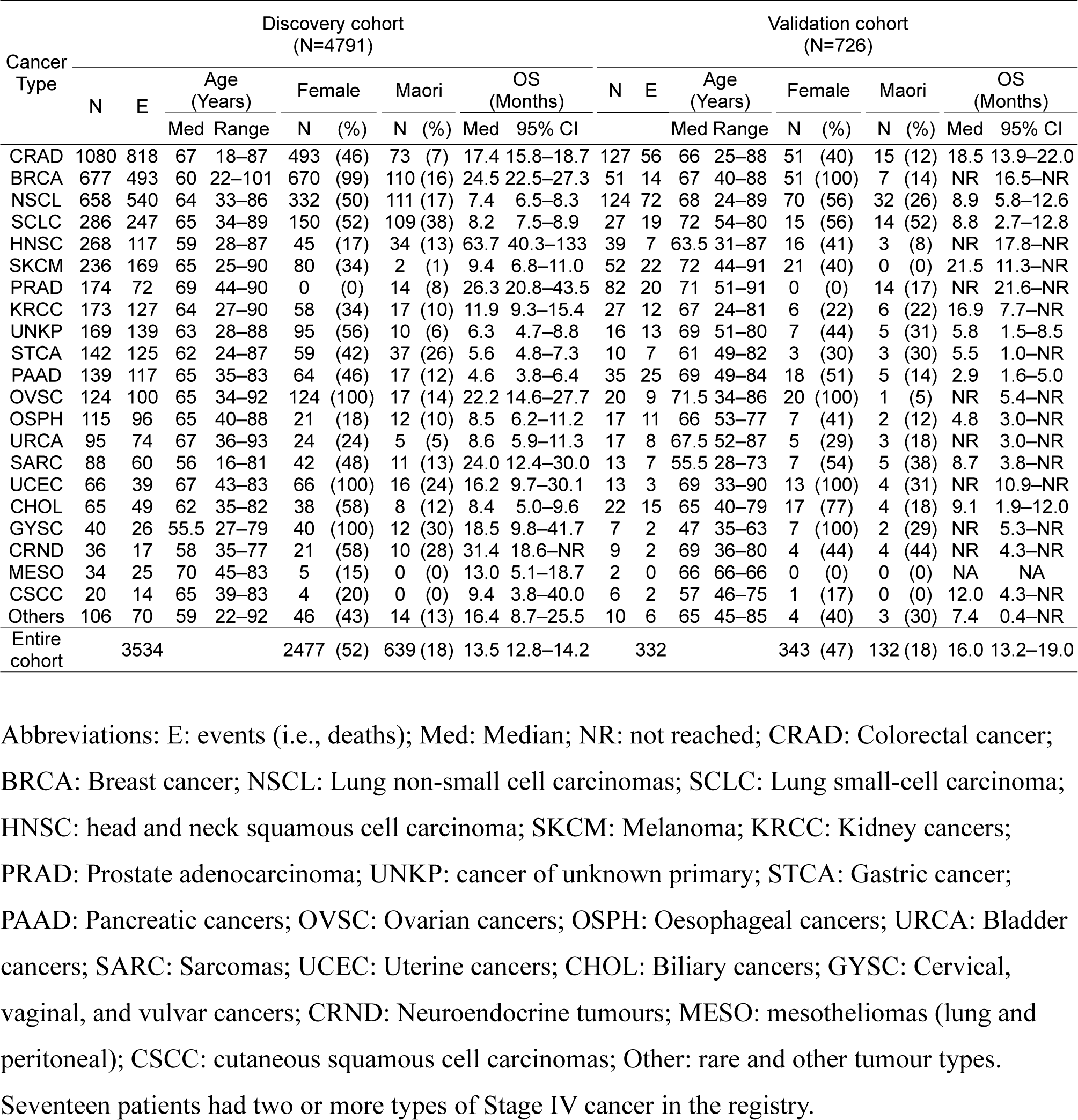
Characteristics of patients in the discovery and validation cohorts by cancer type

A text corpus was first constructed by extracting narratives from correspondence at the initial consultation. We retrieved 349262 EMR letters authored by 115 clinicians, and texts were then cleaned with patient identifiers removed (Table S1). Motifs (text words, short text fragments, and patterns) were extracted by TEPAPA, and mentions of each motif were correlated to normalised survival (nOS; defined as the ratio of survival duration in days to median OS for a given cancer type, Figure S2). TEPAPA identified 5855 unigrams (i.e., words or tokens delimited by spaces or punctuation), with at least 23 (0.5%) mentions in initial consultation correspondence. A false discovery rate (FDR) of 0.001 (Type 1 error rate, α=2.2×10^−6^) enriched this to 120 unigrams; the most significant (Figure 2A) were “*palliative*” (HR 1.57, p=6.4×10^−42^), “*Sevredol*” (a brand of morphine; HR 1.81, p=3.4×10^−26^), and “*hospice*” (HR 2.02, p=1.7×10^−25^).

**Figure 1.**
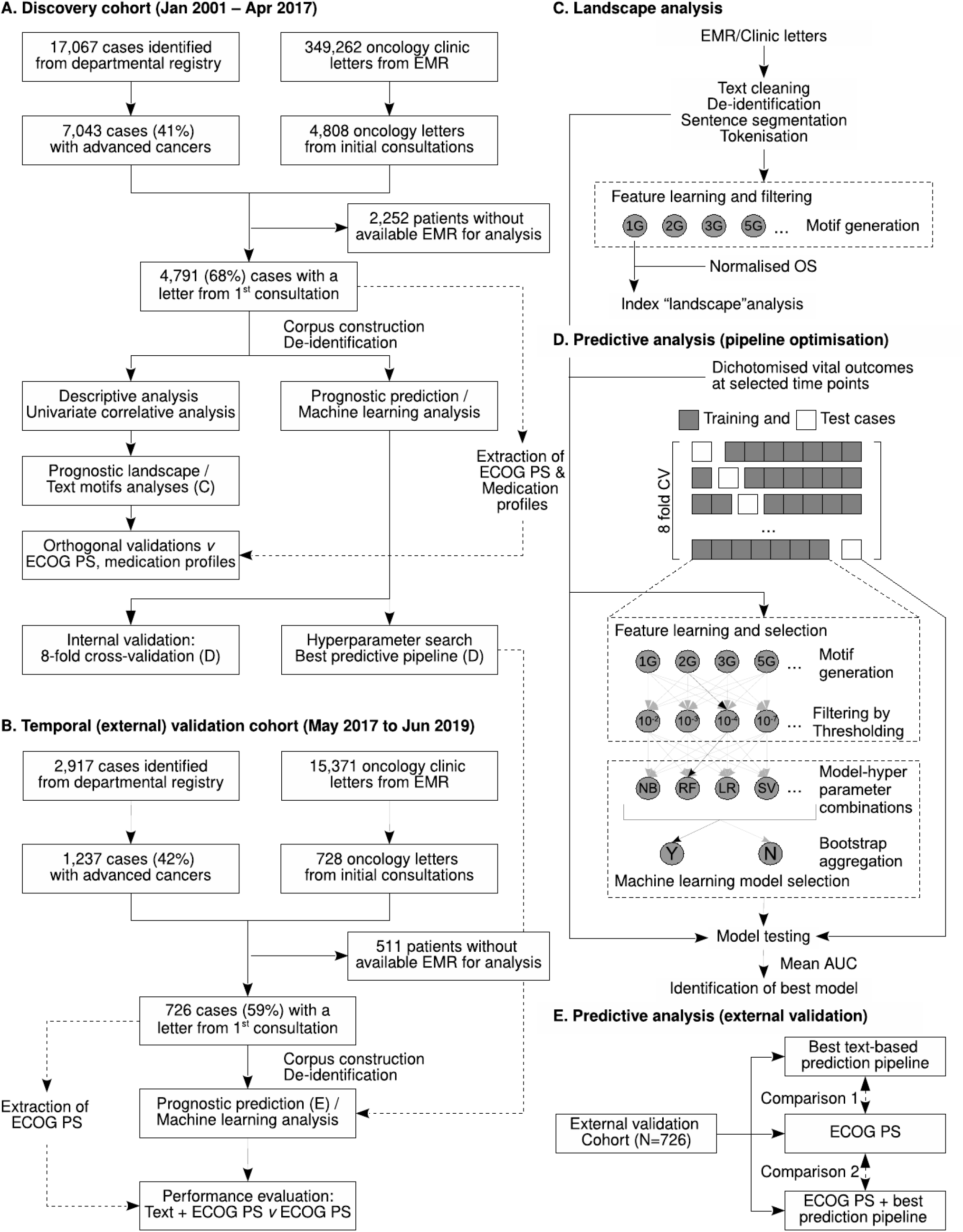
Flow diagram of data analysis. Flow diagram outlining the analytic process of discovery (A) and validation (B) cohorts. Transformation of electronic medical records for landscape analysis (C), predictive pipeline optimisation (D) and external validation (E) are also illustrated. Abbreviation: EMR: Electronic medical record. ECOG PS: Eastern Cooperative Oncology Group Performance Status score. OS: overall survival. AUC: area under receiver operating characteristic (ROC) curve. CV: cross-validation.

**Figure 2.**
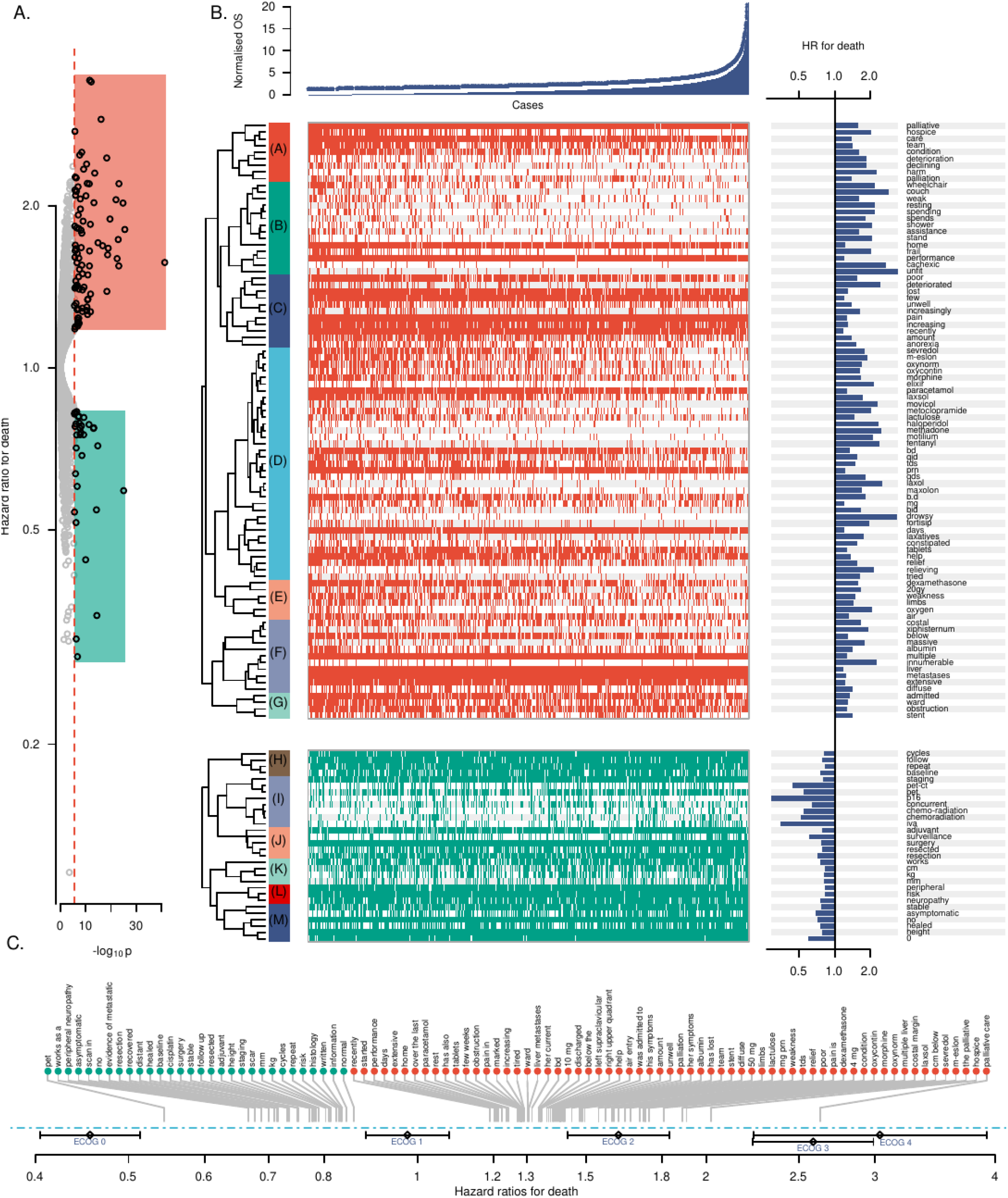
Landscape of cancer type-agnostic prognostic factors in patients with advanced cancers. A. Volcano plot showing univariate associations between the unigrams extracted from clinic letters with normalised OS. Y-axis indicates −log_10_ *p* (log-rank test). The perforated line indicates significance level at FDR q=0.001, corresponding to α=2.2×10^−6^. The complete list of unigrams is shown in Supplementary Data S1. B. (Top) swimmer plot showing survival duration of 4,791 cases. Dots above the bar graph indicate that the case was deceased at censoring date. (Middle and bottom panels) heat maps showing the distribution of mention of a particular word in letters from the first oncology consultation. Clustering of unigrams was carried out by Euclidean distance between 20-dimension embedded text vectors, trained using Word2Vec skip-gram model on the entire available EMR. Clusters are assigned by predominant meanings: (A) Palliative care referral, (B) Mobility and functional status, (C) Symptoms and dynamics, (D) Medications, (E) Brain metastases and treatments, (F) Extensive hepatic metastases, (G) Hospitalization and visceral obstructions, (H) Chemotherapy recommendation, (I) Chemoradiothearpy for HNSCC, (J) Mention of adjuvant treatment, (K) Mentions of “word”, “cm”, and “kg”, (L) Mention of peripheral neuropathy, (M) Asymptomatic Patients. Group K is no longer statistically significant associated with survival in the multivariate logistic regression analysis (Table S3). C. List of *n*-grams (up to 9-grams) that are significantly associated with survival (also see Dataset S3). The average HR for death of ECOG performance status (and 95% confidence interval) to normalised survival are also shown. Abbreviations: HR: Hazard ratio. OS: overall survival. ECOG: Eastern Cooperative Oncology Group performance status score.

To identify concepts represented by extracted motifs, we performed unsupervised clustering of unigrams associated with survival (at FDR=0.001) according to distributional similarity in the EMR. A Word2Vec skip-gram model [15] was trained using the text from 349262 letters to create a 20-dimension embedded vector for each unigram (Supplementary Dataset S2). Using binary hierarchical clustering with Euclidean distance and complete linkage, words were collated into interpretable concept clusters (Figure 2B and Table S3). Clusters of descriptors negatively associated with survival included those relating to palliative care referral (Cluster A, HR for death for each unigram mentioned in the narrative 1.33, *p* = 2.1×10^−67^), patient mobility or functional status (Cluster B, HR 1.32, *p* = 3.0×10^−54^), cancer-related symptoms (Cluster C, HR 1.32, *p* = 7.4×10^−58^), medications including opiates, antiemetics, and laxatives (Cluster D, HR 1.16, *p* = 3.6×10^−50^), brain metastases (Cluster E, HR 1.31, *p* = 6.1×10^−23^), hepatic metastases (Cluster F, HR 1.16, *p* = 1.6×10^−37^), and hospitalization or visceral obstruction (Cluster G, HR 1.24, *p* = 5.3×10^−20^). In cluster B, words seldom used in performance assessments such as “*wheelchair*” (HR 2.12, *p* = 6.9×10^−24^) and “*couch*” (HR 2.90, *p* = 7.4×10^−17^) were often identified.

Descriptors associated with improved survival included chemotherapy-related text (Cluster H, HR 0.83, *p* = 2.5×10^−20^), asymptomatic patients at initial consultation (Cluster M, HR 0.83, *p* = 3.5×10^−27^) and surgical resection, adjuvant therapy, or peripheral neuropathy (Groups J and L). Differences in survival could partly be attributed to lead-time bias in surveillance-detected cases where recurrent, smaller-volume disease is detected earlier (Text S4). In Group I, descriptors associated with HNSCC were suggested by text mining; longer survival is likely attributable in part to 187 of 264 evaluable HNSCC patients (71%) being treated with curative-intent chemoradiotherapy, with longer median OS (120 months, 95% CI: 53.0–not reached) relative to patients treated with palliative intent (25 months, 16.9–40.3, HR 0.52, p=7×10^−4^, Wald test).

As functional status assessment — an accepted standard in prognostication across cancer types [16,17] — was rediscovered by text mining (Clusters B, C, and M), we orthogonally quantified the prognostic capability of Eastern Cooperative Oncology Group (ECOG) scores for patient performance status (PS). In a manual chart review, PS scores were documented by oncologists in 2210 (46%) of the study cohort. ECOG PS was associated with both OS and nOS (both p<0.001, log-rank test, Text S5). Mentions of “*ECOG <number>*” had a comparable HR for ECOG 1-3 (Text S6). Text motifs in clinic letters that are considered “ECOG equivalent” are presented in Figure 2C.

Given that text descriptors describing medication-related concepts were associated with survival (Group D), we examined if supportive and other non-antineoplastic medications retained or prescribed at initial consultation enhanced prognostication. We matched patients’ medications to 4970 World Health Organisation (WHO) Anatomical Therapeutic Chemical (ATC) categories [18]: 4523 of 4791 patients (94%) had medication sections mentioned. A median of 3 ATC codes per patient was identified (interquartile range: 1-5, maximum 16, Figure S7). Penalised regression identified 16 ATC categories associated with survival (Table S8). Survival was significantly associated with analgesics (Table S8), with ATC codes N07BC02 (methadone, HR 1.75, 95% CI: 1.31–2.34) and N02AB03 (fentanyl, HR 1.76, 95% CI: 1.24–2.50) denoting poorest prognosis.

To determine if text motifs from EMR can improve prediction of short-to medium-term mortality, we examined supervised machine learning to build “white box” predictive models. This differs from previous validated machine learning methods of cancer mortality prediction using registry data [19-23] or clinical and physiological variables [24,25]. Classification based on clinical narratives was studied previously to augment prediction of in-hospital mortality in intensive care units [26]. Using “bag-of-motifs” (i.e., presence of term in letter) as features, machine learning models were trained to predict vital outcome at time intervals measured from the beginning of a patient’s treatment journey (every 2 weeks until week 12, then every 4 weeks until week 20, and at 26, 39, and 52 weeks). In 3543 (74%) cases in the discovery cohort where survival duration was determined at the censor date, hyperparameter tuning with exhaustive grid search was conducted by 8-fold cross-validation to assess in-sample performance (Table S9), thus identifying the most accurate classification pipeline, including both feature and machine learning. The random forest model with high bootstrap aggregation (bagging) iterations was the best-performing algorithm by areas under the ROC curve (AUC) in 10 of 13 timepoints (Figure 3A). In cross-validation analysis, the best machine learning model outperformed ECOG PS scores as predictors across all time points (mean AUC improvement: 0.014, *p* = 0.0064, chi-square test, Figure 3B). An additional bagging step (10 iteration with full size training set) on the best classifier further improved performance (mean AUC improvement 0.011, *p* = 0.018, Figure 3C).

**Figure 3.**
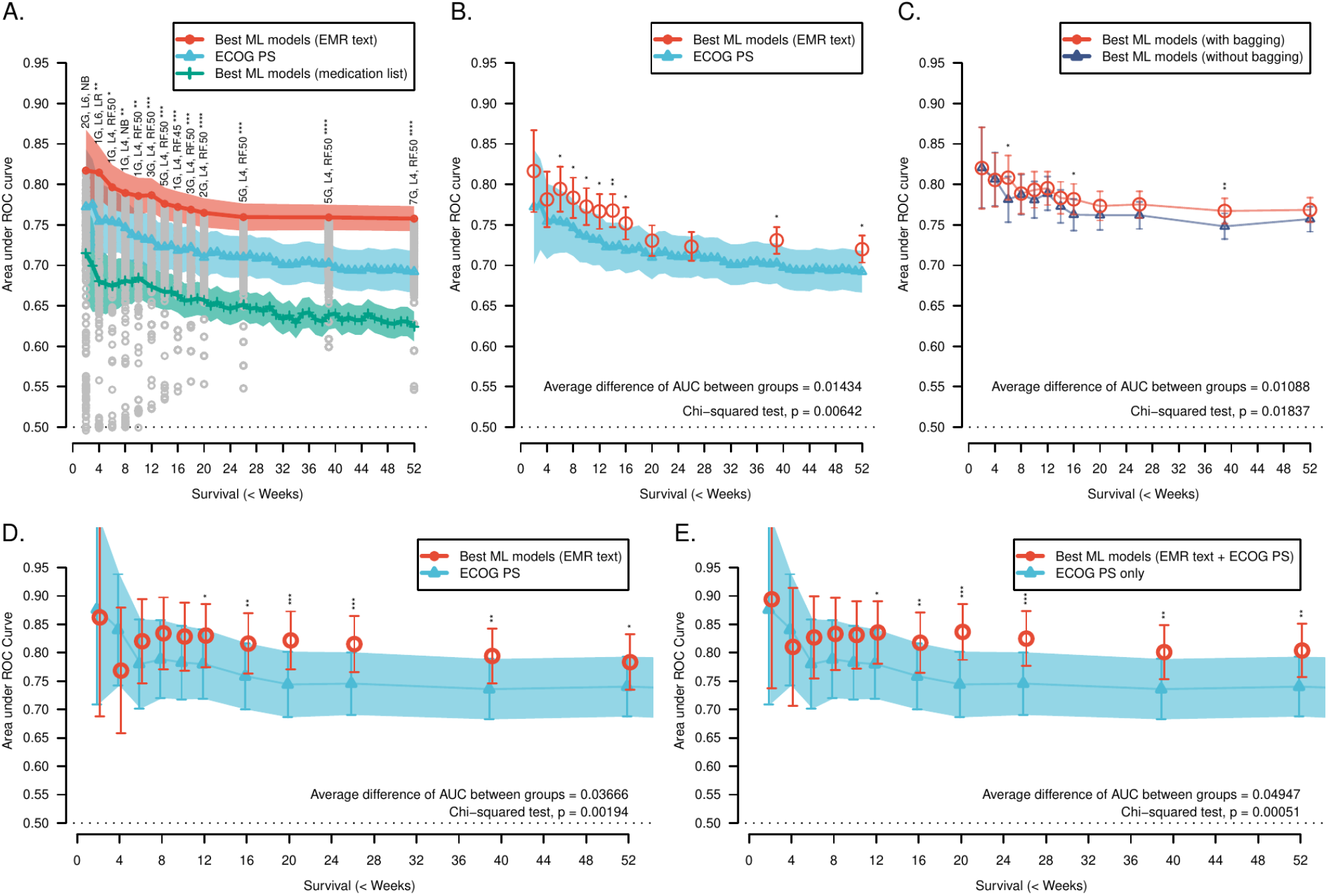
Identification of the optimal machine learning pipelines and external validation results. A. Results from cross-validation (CV) procedure for identifying the most predictive machine learning model using grid-based hyperparameter search. Classification performance was estimated by mean area under ROC curve (AUC). Random Forest (with a high number of iterations) consistently outperformed other classifiers in the task. The length of *n*-gram motifs was not associated with AUC, although the feature filtering threshold was an important determinant (p<0.001). The optimal threshold was between −log_10_ α=4-6. Corresponding AUC of predictors using medication profiles and ECOG scores are also shown for comparison. Abbreviations: *n*G: *n*-gram motifs, L4/L6: filtering threshold for selecting unigrams with p<10^−4^ and 10^−6^ respectively; LR: Logistic regression; NB: Naive Bayes; RF*n*: Random forest with *n* bagging iterations. B. CV (8-fold) results of the best prognostic pipeline (i.e., feature learning with ML model) for survival prediction at selected time points. Overall, machine learning models outperforms ECOG PS in survival prediction. Using clinical narratives, the machine learning-based method predicts survival of patients at 2, 6, 12, 26, 52, and 80 weeks was predicted with AUC of 0.82, 0.80, 0.77, 0.72, 0.72, and 0.76 in the discovery cohort; machine learning model was non-inferior to ECOG scoring at all time thresholds. C. CV results examining the performance of the best classifiers with or without bagging (with 10 bootstrap iterations with 100% sample size): bagging of top performing classifiers further improved classifier performance significantly. D & E. The performance of best performing pipeline in the external validation cohort of 726 patients. The best pipeline (including both feature learning, supervised classifier selection, and hyperparameter tuning) was identified by training machine learning models using oncologists’ narratives from the discovery cohort. Overall, using clinical text with or without the inclusion of ECOG performance status in a machine learning model significantly improved survival prediction over ECOG scoring alone. Mean AUCs of combined ECOG and text model were >0.80 at all assessed time points in the group with ECOG with text prediction. Legend: P<0.10 (*), <0.05(**), and <0.01 (***) respectively. Shaded areas and vertical bars indicate 95% confidence intervals.

External validity of text-based prognostic models was examined with the best machine learning models in a temporal (validation) cohort of 1237 cases Stage IV cancers (of 2917 patients) from May 2017 to Jun 2019; 726 patients had available correspondence at the initial oncology visit. At a median follow-up of 15.8 months (95% CI: 14.6–17.3), OS was comparable across cancer types except melanoma (median OS: 21.5 vs. 9.4mo, HR 0.58, *p* = 0.02, Figure S10). Across all timepoints, text-based classifier was superior to ECOG PS in survival prediction (mean AUC improvement 0.037, *p* = 0.0019, chi-square test, Figure 3D and Table S11). When combining text-based features with ECOG PS scores in machine learning models, the hybrid classifiers overall performance was superior to using ECOG PS alone (mean AUC improvement 0.049, *p* = 0.0005, Figure 3E and Table S12), supporting a prognostic classification scheme by re-purposing clinical narrative in survival prediction.

In summary, the above EMR analysis identified novel prognostic factors in advanced cancers, and demonstrates how unpatterned clinical text stored in EMR may be translated into useful concepts. Our comparative predictive analyses illustrate how clinical free-text can augment prognostic accuracy over traditional predictive approaches, potentially improving the monitoring and tailoring of palliative therapy. Machine-analysable text in EMR creates new opportunities to digitalise clinical assessments, improve cancer care outcomes, and perhaps also identify patients for trials.

## Supporting information

Supplemental data set

## Data Availability

The original data set is not available, with the exception of summarised and non-reidentifiable datasets supplied as supplementary text. Computer code associated with this manuscript is released as open-source software and is freely available.

## Acknowledgements

FPL was supported by Shine Translational Fellowship 2016, Garvan Institute of Medical Research. This project is partly supported by a research project grant of Waikato Research Foundation 2018. This work has been previously presented in abstract and poster forms at ASCO Breakthrough Asia meeting 2019, Bangkok, Thailand [27]. The authors thank Dr. Ian Kennedy for assistance in data extraction and Dr. Alvin Tan for comments. FPL and RE acknowledge the support from the Wolf family. The authors declare no conflict of interest. This study was approved by Northern Health & Disability Ethics Committee, New Zealand (#16/STH/251).

## ONLINE METHODS

### Study type and cohort characteristics

This retrospective cohort study was conducted at a tertiary regional cancer centre, including three peripheral small urban or rural clinics, in New Zealand, serving a total population of 700,000. Demographic data of each patient was extracted from the departmental registry, including age, gender, survival duration, ethnicity (Maori vs Non-Maori), cancer type, and stage. Vital status and date of death were also retrieved from a departmental registry, secondarily derived from the Ministry of Health of New Zealand. All patients diagnosed with Stage IV solid tumours at the first consultation from January 2001 to April 2017, were included. Patients with haematological malignancies, including leukaemia and lymphomas, or survivable cancers, were excluded from analysis. Survival data for the discovery cohort was censored on 18 April 2017.

### Descriptive and survival analysis

For descriptive analysis, cases were broadly grouped into 21 categories with a group of “other” which contain histology types that are otherwise not classifiable. In the primary landscape analysis (see below), median survival by cancer type was assessed by Kaplan-Meier analysis. The median time of follow-up was assessed by reverse Kaplan-Meier method [28].

Considering the diverse survival across different cancer types, we calculated normalised survival (nOS) to allow the discovery of prognostic covariates in a cancer type-independent manner. For each patient, the normalised survival was calculated such that:

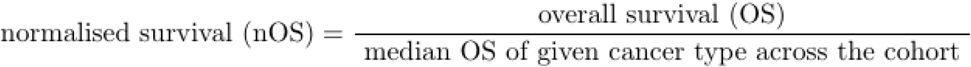

where the length of survival of each patient is adjusted by tumour type in our cohort (Figure S2). The nOS loosely measures the “stage” of patient’s journey with respect to the underlying cancer type.

### Construction of corpus for text mining

The corresponding clinical letter of each patient was accessed from the departmental audit database. The clinical narrative from the first specialist consultation was extracted. To ensure each patient is represented identically and independently distributed, we included only the letter from first consultation per patient, also as to avoid selection bias towards patients with a higher number of clinical documentations (i.e., patients with more visits to the oncology clinic).

#### Data cleaning

A custom script was developed to automatically redact identifiable information, including names of patient, clinicians, addresses, date of birth, date of consultation, healthcare facilities, affiliation of healthcare professionals (including oncologists, primary care provider, and associated specialists) and they are replaced by tokens. *Sentence segmentation* was then performed, and text was folded into lower case. Tokenisation was performed by delimiting a string of text by white spaces and punctuations, as outlined in our previous work [13].

### Informatics analysis

#### Extraction of unigrams associated with survival

TEPAPA was applied to examine the association between a motif (unigram and other *n*-grams where *n ≥* 2) with nOS. The *n*-gram text motifs were identified by iterating through the entire sequence of text using an exhaustive search method as previously reported [13]. In the index analysis, unigrams with mentions of ≥ 0.5% of the sample size (i.e. ≥ 23 cases per patient for the index analysis 4,791 patients) were included in the survival analysis; Kaplan-Meier method with univariate hypothesis testing was performed. The prognostic value of each unigram was analysed with hazard ratio (HR) for death between cases where the motif is mentioned to certain phases to survival identified was calculated log-rank test:

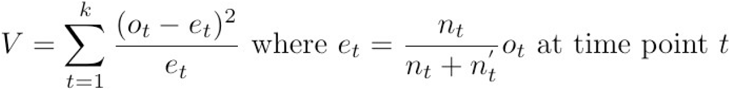

where V is the log-rank statistic, *o*_*t*_ is an observed event at time point *t, e*_*t*_ is the expected events at time point *t*, and *n*_*t*_ and *n*_*t*_*’* is the numbers at risk where a given text motif was mentioned or not mentioned respectively. The type I error rate was estimated by chi-squared test with one degree of freedom where. The HR for death λ between the groups (i.e., whether a motif was mentioned vs. not-mentioned) is estimated by Bernstein method [29], where:

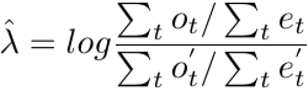

Significance of each unigram was filtered by false discovery rate (FDR) using Benjamini-Yekutieli procedure (BY-FDR) [30] where the null hypothesis is rejected if *p* < *P*_(*k*)_ :

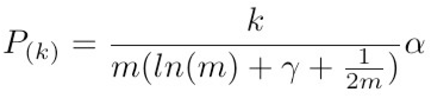

#### Unsupervised clustering of unigrams

In the index analysis, we trained a Word2Vec skip-gram model to represent each unigram as a 20-dimension vector to measure distributional similarity between unigrams within contextual meanings [15]. The skip-gram model was trained by using the entire cohort of 349,262 clinic letters in 17,067 patients, with parameters were set with a skip-length of 16 tokens, 25 tokens for negative sampling, and a sampling threshold set at 10^−4^, using 10 training iterations. Skip-gram was selected given that it provides a better interpretation compared to the Continuous Bag of Words model or by occurrence within EMR. Binary hierarchical clustering was then performed, using Euclidean distance between the embedded vectors with complete linkage. Thresholding of clusters was manually conducted by the lead author (FL).

#### N-gram analysis

The threshold of was applied FDR at q=0.001 was performed in N-gram vectors. Hazard ratios of the prognostic value of a motif was also reported as per unigram analysis above.

### Orthogonal validations

Numerical score of performance status (using keyword “*WHO”, “Zubord”*, or *“Karnofsky/KPS”*) at the time of first consultation was extracted from the narrative. Karnofsky score was converted to ECOG score according to the functional scale by Oken et al [3]. Kaplan-Meier analysis on nOS for patients was tested by Multivariate Cox’s regression, and HR for death was estimated.

#### Medication profiles

The list of medication of each patient documented by oncologists at the end of first oncology consultation was recorded (included both routine regular medications and prescribed at the consultation). Medication allergies, alternative medicines (e.g., herbal remedies), and chemotherapeutic agents were excluded from analysis. Trade names were mapped to a list of ingredients using a database extracted from Medsafe New Zealand [31]. The ingredients were mapped to WHO Anatomical Therapeutic Category [18]; ingredients with less than 20 mentions were excluded from analysis. Descriptive analysis was carried out by enumerating the frequency of medication mentions. Least Absolute Shrinkage and Selection Operator (LASSO) Cox’s regression was performed to identify the covariates using glmnet package [32], using the minimum lambda value identified by cross-validation for regularisation of variables. The mapping of ATC category is available in the Supplementary Material.

### Machine learning and predictive analysis

#### Construction of machine learning models for survival prediction using clinical text

The entire discovery cohort, where survival is known at the censor date, was dichotomised into groups by (absolute) duration of survival. Patients were dichotomised into clinically-significant intervals listed in the main text. Supervised machine learning algorithms were used to examine whether the text motifs extracted by TEPAPA can predict vital status (alive vs. death) at different time points. Different classes of machine learning algorithms (naive Bayesian, logistic regression, alternating decision trees, support vector machines with polynomial kernels, and random Forest) were evaluated.

#### Hyperparameter tuning and identification of optimal machine learning models

The optimal hyperparameters of the pipeline was identified by grid search: the optimal combination of feature and machine learning pipeline at each interval was assessed by AUC using 8-fold cross-validation. The combinations of hyperparameters examined in this study is listed in Table S9. Previously, bootstrap aggregated (“bagged”) models were shown to reduce variance thus improving accuracy [33], and we have previously reported that they outperform the non-aggregated counterparts in a breast cancer cohort [34]. To reduce intensity of computational resources, bootstrap aggregated models were further performed only to the best machine learning models where optimal combinations of hyperparameters were determined. At each timepoint, the best pipeline was identified as the one classifier that achieved the highest mean AUC across all folds in the cross-validation.

#### Estimation of predictive performance of best pipeline by cross-validation

The out-of-sample performance of classifier was estimated by a separate cross-validation run with, where a separate test fold was held out, independent from feature selection and model training steps. In each fold, training data was first used to identify the best prediction pipelines (both feature and machine learning models), optimised by features extracted by TEPAPA algorithm, with variation in the hyper-parameters as per hyperparameter search. The mean AUC across all folds was obtained to estimate the out-of-sample performance.

#### External (Temporal) validation

Predictive performance of machine learning model trained by clinical narratives, with or without incorporation of ECOG PS score as a covariate, was compared to ECOG PS score alone for predicting vital outcome in a temporal cohort at the same study site. The temporal cohort was identified from consecutive cases over the following 25 months and were screened at the same study site. Using the optimised pipeline identified above, performance of classifier in predicting short-to mid-term mortality was assessed by AUC in survival classification at 11-point time intervals (at 2, 4, 6, 8, 10, 12, 16, 20, 26, 39, and 52 weeks, respectively) reflecting practical timepoints in clinical decision making. The models derived from the discovery cohort were used to predict vital status at each time point. Text corpus was obtained and processed identically to the procedure described in the discovery cohort.

Two comparisons were performed (comparison 1: Machine learning models trained with clinical narratives vs ECOG, and comparison 2: machine learning models trained with clinical narratives with ECOG v ECOG). In Comparison 2, predictive text motifs were first identified by the feature learning algorithm and ECOG PS scores was incorporated as a covariate. Patients without documented ECOG performance status are assigned a missing class. In both comparisons, the mean differences in AUC between groups across all time points were calculated; statistical hypothesis testing was conducted by Chi-square goodness-of-fit test with variance of AUC estimated by Hanley-McNeil method [35]. Pairwise comparison of AUC between groups was conducted by using Z-tests for individual time points.

#### Software

Waikato Environment for Knowledge Analysis (WEKA) version 3.6.6 [36] and R environment for statistical computing were used (Naive Bayes) for training and evaluating machine learning models. Both TEPAPA and framework for building and evaluating predictive machine learning pipelines are available at http://tepapadiscoverer.org/.

## Notes

### Competing Interest Statement

The authors have declared no competing interest.

### Funding Statement

FPL was supported by Shine Translational Fellowship 2016, Garvan Institute of Medical Research. This project was partly supported by a research project grant of Waikato Research Foundation 2018. FPL and RE acknowledge the support from the Wolf family.

### Author Declarations

This study was approved by Northern Health & Disability Ethics Committee, New Zealand (#16/STH/251).

