## Supplemental text for "Repurposing digitised clinical narratives to discover prognostic factors and predict survival in patients with advanced cancer"

| Item | Description |
|---|---|
| **Table S1** | Characteristics of text corpus used in prognostic factor discovery. |
| **Figure S2** | Kaplan-Meier survival curves of normalised versus non-normalised survival by cancer type |
| **Table S3** | Prognostic cluster identified in the index analysis |
| **Text S4** | Survival characteristics of patients who has had previous curable colorectal cancers. |
| **Text S5** | Hazard Ratios for death by ECOG performance status. |
| **Text S6** | Comparison between documented ECOG PS and extraction of ECOG mentions in oncologists' letters from the first clinic visits |
| **Figure S7** | Distribution of number of non-anti-cancer drugs retained or prescribed by oncologists at the first oncology assessments. |
| **Text S8** | Prognostic significance of non-cancer medications retained or prescribed at the first oncology consultation significantly associated with survival. |
| **Table S9** | Hyperparameters of feature learning, selection, classifier, and examined in the classification task. |
| **Figure S10** | Overall survival of patients in the discovery and validation cohorts by cancer type |
| **Table S11** | Comparison of prognostic accuracy of ECOG performance status versus clinical text in predicting short to mid-term mortality in external validation cohort. |
| **Table S12** | Comparison of prognostic accuracy of ECOG performance status with and without clinical text in predicting short to mid-term mortality in patients with advanced cancers in the external validation cohort. |
| **Text S13** | Patient with multiple diagnoses of advanced cancers in the validation cohort |
| **Text S14** | Additional discussions |
| **Figure S15** | Principal component analysis of the 20-dimension vectors of the unigrams discovered in the index analysis. |
| **Figure S16** | Overall survivals in Maori and non-Maori patients of entire cohort |
| **Figure S17** | Overall survivals in Maori and non-Maori patients of entire cohort by cancer types |
| **Table S18** | STrengthening the Reporting of OBservational studies in Epidemiology (STROBE) |
| **Table S19** | REporting recommendations for tumour MARKer prognostic studies (REMARK) statement |

**Supplementary Table S1 – Characteristics of text corpus used in prognostic factor discovery.**

| Characteristic | Discovery cohort (N=4,791) (Jan 2001 – Apr 2017) | | Validation cohort (N=726) (May 2017 – Jun 2019) | | P |
|---|---|---|---|---|---|
| Duration (Months) | 197 | | 25 | | |
| Total correspondences * | 349,263 | (1,773 per month) | 15,731 | (629 per month) | <0.001 [a] |
| Unique patient files | 4,791 | (24.4 per month) | 726 | (29.0 per month) | 0.47 [a] |
| Unique authors * | 115 | | 33 | | 0.007 [a] |
| - Oncologists | 4,747 | (99%) | 722 | (99%) | 0.97 [a] |
| - Specialist registrars * | 1,298 | (27%) | 272 | (37%) | <0.001 [a] |
| Files per author (Median) | 8 | IQR: 3-29<br>Range: 1-2,048 | 16 | IQR: 8-47<br>Range: 2-144 | 0.052 [b] |
| Paragraphs per file * | 32.5 | (95% CI: 10-55) | 21.4 | (95% CI: 5.6-36.8) | <0.001 [b] |
| Words per file | 701 | (95% CI: 147-1,256) | 712 | (95% CI: 236-1,190) | 0.08 [c] |
| Bytes per file | 4,744.2 | (95% CI: 1,337-8,152) | 4,644.5 | (95% CI: 1,758-7,532) | 0.29 [c] |

No significant difference between the number of unique patients per month comparing the discovery and validation periods was found. Nearly all letters were authored by consultant oncologists with or without specialist registrars. There was no difference between the number of words, files per authors, and length of file. The difference in paragraphs per file reflects changes in document structure between the two periods. Notes: [a] Chi-square test with 1-degree of freedom (vs number of months). [b] Kolmogorov-Smirnov test to examine difference in distribution number of files per authors between the discovery and validation cohorts. [c] Student's *t*-test.

**Supplementary Figure S2 – Kaplan-Meier survival curves of normalised versus non-normalised survival by cancer type**

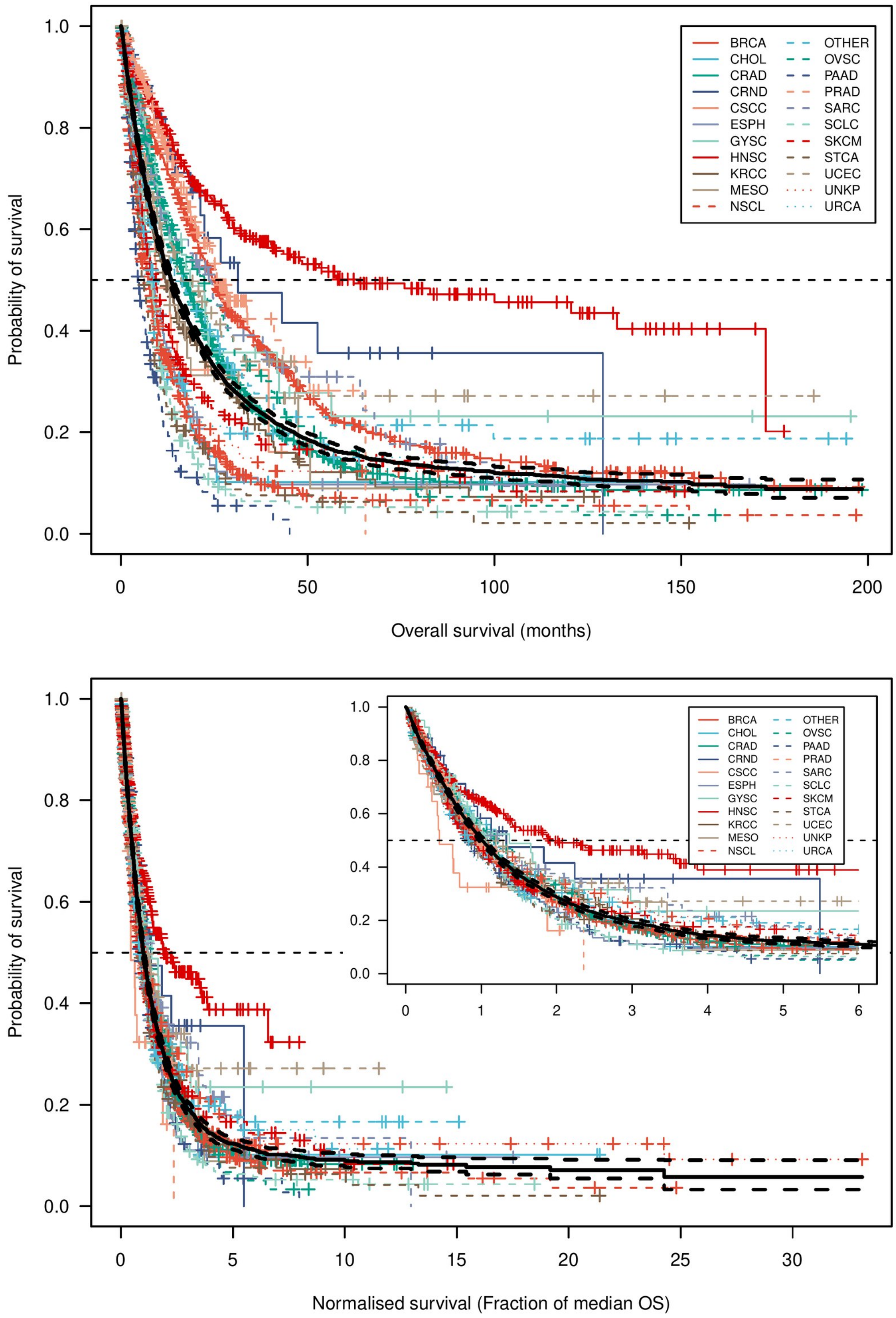


Overall survival (Top) versus normalised survival (Bottom) demonstrating the effect adjustment by cancer type. The bold black line indicates OS (top) and nOS (bottom) of the overall population respectively.

**Supplementary Table S3 – Prognostic cluster identified in the index analysis**

| Semantic cluster | Mentioned * | | Univariate model | | | Multivariate model | | |
|---|---|---|---|---|---|---|---|---|
| | N | (%) | HR | 95% CI | P | HR | 95% CI | P |
| *Poor prognostic groups* | | | | | | | | |
| (A) Palliative care referral | 3,060 | (64) | 1.33 | 1.29-1.37 | $2.1\times10^{-67}$ | 1.17 | 1.13-1.21 | $<2\times10^{-16}$ |
| (B) Mobility and functional status | 2,663 | (56) | 1.32 | 1.27-1.37 | $3.0\times10^{-54}$ | 1.14 | 1.10-1.18 | $4.5\times10^{-12}$ |
| (C) Symptoms and dynamics | 4,239 | (88) | 1.21 | 1.18-1.24 | $7.4\times10^{-58}$ | 1.11 | 1.08-1.14 | $1.3\times10^{-15}$ |
| (D) Medications | 3,150 | (66) | 1.16 | 1.14-1.19 | $3.6\times10^{-50}$ | 1.07 | 1.04-1.09 | $2.1\times10^{-8}$ |
| (E) Brain metastases | 1,159 | (24) | 1.31 | 1.24-1.38 | $6.1\times10^{-23}$ | 1.12 | 1.06-1.18 | $6.5\times10^{-5}$ |
| (F) Hepatic metastasis | 3,996 | (83) | 1.16 | 1.13-1.19 | $1.6\times10^{-37}$ | 1.10 | 1.08-1.13 | $<2\times10^{-16}$ |
| (G) Hospitalization; (visceral) obstructions | 1,495 | (31) | 1.24 | 1.18-1.30 | $5.3\times10^{-20}$ | 1.06 | 1.01-1.11 | 0.012 |
| *Good prognostic groups* | | | | | | | | |
| (H) Chemotherapy recommendation | 2,743 | (57) | 0.83 | 0.79-0.86 | $2.5\times10^{-20}$ | 0.87 | 0.83-0.91 | $6.7\times10^{-11}$ |
| (I) Chemoradiotherapy for HNSCC | 1,150 | (24) | 0.76 | 0.71-0.81 | $1.3\times10^{-16}$ | 0.85 | 0.79-0.91 | $1.2\times10^{-6}$ |
| (J) Mention of adjuvant treatment | 2,654 | (55) | 0.83 | 0.80-0.86 | $2.1\times10^{-30}$ | 0.92 | 0.89-0.95 | $3.2\times10^{-7}$ |
| (K) Mentions of “works”, “cm”, and “kg” | 1,121 | (23) | 0.92 | 0.87-0.97 | $9.4\times10^{-4}$ | 0.96 | 0.91-1.01 | 0.144 |
| (L) Mention of peripheral neuropathy | 2,327 | (49) | 0.87 | 0.84-0.91 | $1.5\times10^{-13}$ | 0.89 | 0.86-0.92 | $8.1\times10^{-10}$ |
| (M) Asymptomatic patients | 4,287 | (89) | 0.83 | 0.80-0.86 | $3.5\times10^{-27}$ | 0.88 | 0.85-0.91 | $4.4\times10^{-12}$ |

Prognostic value of semantic clusters grouped by skip-gram vectors at FDR of 0.001. The Hazard Ratio (HR) reported here indicates the average HR for death per unigram (from the cluster) mentioned in the clinic letter. After a multivariate analysis considering all factors, clusters (G) and (K) are no longer statistically significant (at Bonferroni corrected $\alpha=0.0038$). Abbreviation: HNSCC: Head and neck squamous cell carcinoma; HR: hazard ratio for death; Mentioned: number of letters where a unigram in the cluster was mentioned.

**Supplement Text S4 – Survival characteristics of patients who has had previous curable colorectal cancers.**

| Characteristic | | Contrast | | Univariate HR | | Multivariate HR | | |
|---|---|---|---|---|---|---|---|---|
| | N | | N | Estimate | P | Estimate | (95% CI) | P |
| Relapsed diseases | 149 | De novo metastasis | 931 | 0.71 | 0.0010* | 0.717 | (0.58-0.88) | 0.0018* |
| Age (per year) | | | | 1.009 | 0.0027* | 1.009 | (1.003-1.015) | 0.0053* |
| Male sex | 587 | Female | 493 | 0.98 | 0.98 | 1.010 | (0.88-1.16) | 0.89 |
| Ethnicity = Maori | 73 | Non-Maori | 1007 | 0.85 | 0.30 | 0.92 | (0.67-1.14) | 0.58 |
| Site Rectal | 258 | Colon | 822 | 0.79 | 0.0060* | 0.82 | (0.95-0.97) | 0.023* |

In clusters (J) and (L), mentions of "adjuvant" or "peripheral neuropathy" were found to be significantly associated with longer survival in cancer patients. To further examine whether prior curative cancer treatment is also associated survival, we studied whether patients diagnosed with relapsed colon and rectal cancers after curative treatment are associated with longer survival. In the discovery cohort, 149 of 1080 (14%) patients with advanced colorectal cancer patients who had previously been consulted at the index cancer centre. In a multivariate logistic regression analysis, patients who was diagnosed with *de novo* metastatic disease had a shorter median overall survival (OS) of 16.3 months (95% CI: 14.2-17.9), comparing to patients who has had a diagnosis with Stage II (N=8) or Stage III (N=133) diseases (median OS: 24.5 months, 95% CI: 19.0-31.7; p=0.001, Log-rank test). Our data thus suggests that patients who had documented curative treatments of colorectal cancers are associated with a significantly longer survival compared to discovery of *de novo* metastasis (HR 0.72, p=0.0018). While our retrospective data is unable to examine a causative rule, there is a strong likelihood of lead-time bias of patients undergoing surveillance, which resulted in earlier detection of metastatic disease with a lower disease burden.

**Supplementary Text S5 – Hazard Ratios for death by ECOG performance status.**

| | | ECOG PS | N | (%) | HR | (95% CI) |
|---|---|---|---|---|---|---|
| **Discovery cohort** | Overall survival | 0 | 592 | (27) | Reference | |
| | (N=2214) | 1 | 925 | (42) | 1.79 | (1.57-2.04) |
| | | 2 | 416 | (19) | 2.80 | (2.41-3.26) |
| | | 3 | 237 | (11) | 4.74 | (3.99-5.63) |
| | | 4 | 44 | (2) | 5.33 | (3.88-7.32) |
| | Normalised survival | 0 | 590 | (27) | Reference | |
| | (N=2210) | 1 | 923 | (42) | 1.73 | (1.52-1.98) |
| | | 2 | 416 | (19) | 2.64 | (2.28-3.08) |
| | | 3 | 237 | (11) | 4.23 | (3.56-5.03) |
| | | 4 | 44 | (2) | 5.37 | (3.91-7.39) |
| **Validation cohort** | Overall survival | 0 | 95 | (22) | Reference | |
| | (N=439) | 1 | 212 | (48) | 2.00 | (1.23-3.24) |
| | | 2 | 91 | (21) | 4.59 | (2.76-7.61) |
| | | 3 | 33 | (8) | 7.90 | (4.45-14.0) |
| | | 4 | 8 | (2) | 12.4 | (5.19-29.5) |

Stratified Kaplan-Meier analysis of the discovery and validation cohorts by ECOG performance scores in the cases of ECOG status was explicitly documented in the EMR.

**Supplementary Text S6 – Comparison between documented ECOG PS and extraction of ECOG mentions in oncologists' letters from the first clinic visits**

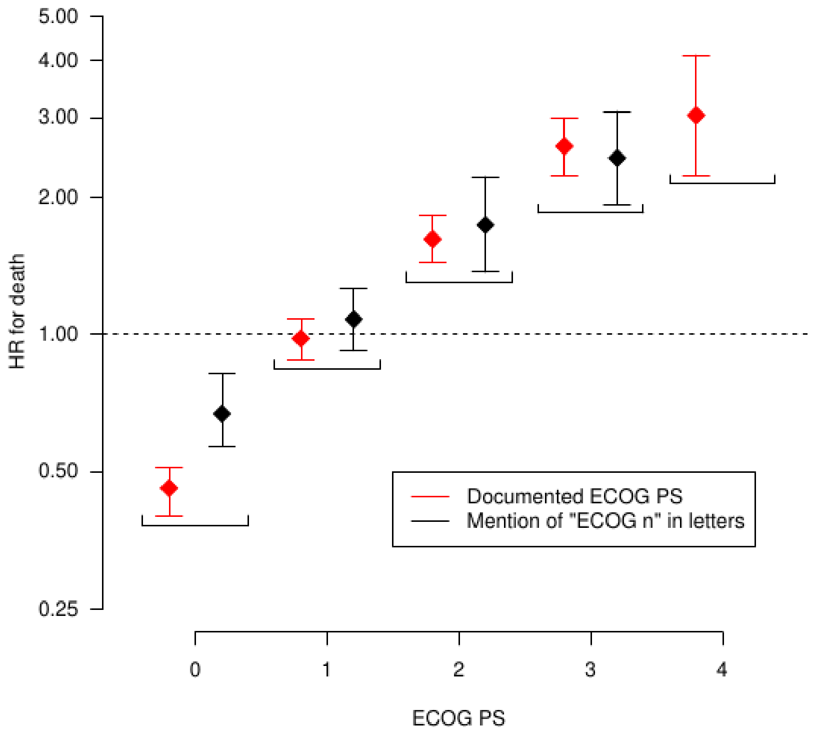


| ECOG PS | Documented ECOG PS (vs the remain group) | | | | Mentions of bi-gram *"ECOG n"* in the text (vs no mentions) | | | |
|---|---|---|---|---|---|---|---|---|
| | N | HR | 95% CI | P | N | HR | 95% CI | P |
| 0 | 590 | 0.46 | (0.40--0.51) | $< 2 \times 10^{-16}$ | 179 | 0.67 | (0.82--0.57) | $3.5 \times 10^{-5}$ |
| 1 | 923 | 0.98 | (0.88--1.08) | 0.64 | 205 | 1.08 | (0.92--1.26) | 0.36 |
| 2 | 416 | 1.62 | (1.44--1.83) | $6.4 \times 10^{-15}$ | 75 | 1.74 | (1.37--2.22) | $7.1 \times 10^{-6}$ |
| 3 | 237 | 2.59 | (2.24--2.99) | $< 2 \times 10^{-16}$ | 69 | 2.44 | (1.93--3.09) | $1.2 \times 10^{-3}$ |
| 4 | 44 | 3.03 | (2.24--4.11) | $8.1 \times 10^{-13}$ | - | - | - | - |

Kaplan-Meier analysis comparing explicitly documented ECOG scores, versus mention of string substring "*ECOG n*", showing that have a comparable hazard ratio for death in ECOG PS of 1-3.

**Supplementary Figure S7 – Distribution of number of non-anti-cancer drugs retained or prescribed by oncologists at the first oncology assessments.**

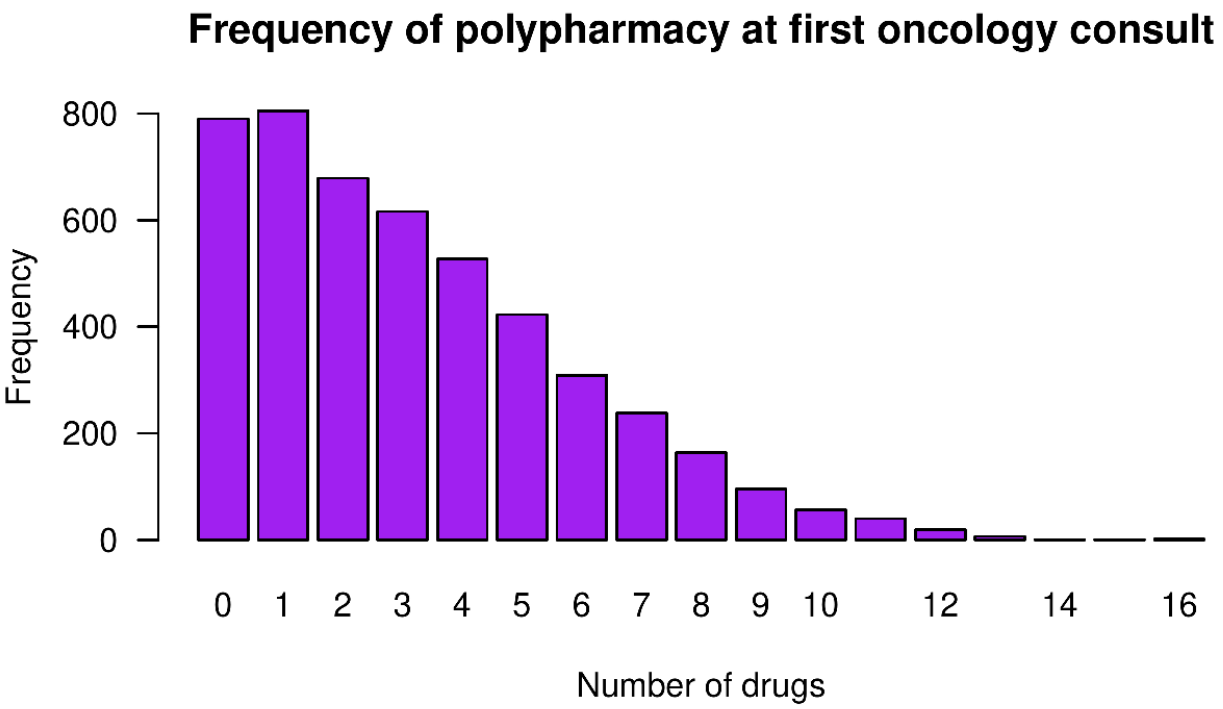


Distribution of number of prescribed medications at the initial oncology consultation. A median of 3 non-cancer drugs per patient (IQR 1-5) was retained from the previous prescription or prescribed by the oncologist after the initial consultation.

**Supplementary Text S8 – Prognostic significance of non-cancer medications retained or prescribed at the first oncology consultation significantly associated with survival.**

| Ingredient | | Frequency (N=4,774) | | HR for death | | |
|---|---|---|---|---|---|---|
| ATC code | Name | N | % | Estimate | 95% CI | P |
| Drugs significantly associated with a poor prognosis | | | | | | |
| N07BC02 | Methadone | 40 | (0.8) | 1.76 | (1.24-2.50) | 0.0017 |
| N02AB03 | Fentanyl | 55 | (1.2) | 1.75 | (1.30-2.34) | 0.00018 |
| N05AA02 | Levomepromazine | 29 | (0.6) | 1.66 | (1.12-2.46) | 0.012 |
| N02AA01 | Morphine | 670 | (14) | 1.60 | (1.45-1.76) | $<1\times10^{-10}$ |
| N05AD01 | Haloperidol | 50 | (1) | 1.56 | (1.15-2.11) | 0.0044 |
| H02AB02 | Dexamethasone | 524 | (11) | 1.49 | (1.34-1.66) | $<1\times10^{-10}$ |
| A04AA01 | Ondansetron | 136 | (3) | 1.42 | (1.17-1.73) | 0.00034 |
| N02AA05 | Oxycodone | 347 | (7) | 1.42 | (1.25-1.61) | $7.1\times10^{-8}$ |
| H02AB07 | Prednisone | 187 | (4) | 1.39 | (1.17-1.65) | 0.00023 |
| C03CA01 | Furosemide | 223 | (5) | 1.37 | (1.17-1.60) | $6.1\times10^{-5}$ |
| A03FA01 | Metoclopramide | 275 | (6) | 1.35 | (1.18-1.56) | $2.2\times10^{-5}$ |
| B01AA03 | Warfarin | 188 | (3) | 1.26 | (1.06-1.48) | 0.0061 |
| N02BE01 | Paracetamol | 1,301 | (27) | 1.12 | (1.04-1.21) | 0.0033 |
| Drugs significantly associated with a good prognosis | | | | | | |
| C10AA05 | Atorvastatin | 278 | (6) | 0.74 | (0.63-0.88) | 0.00056 |
| R03BA02 | Budesonide | 95 | (2) | 0.66 | (0.51-0.86) | 0.0017 |
| C08CA01 | Amlodipine | 56 | (1) | 0.61 | (0.41-0.89) | 0.011 |

A multivariate logistic regression analysis using covariates selected by LASSO regression (see methods). Antineoplastic medications (chemotherapeutic agents, biological therapies, and targeted therapies) were excluded from the analysis.

**Supplementary Table S9 – Hyperparameters of feature learning, selection, classifier, and examined in the classification task.**

| | | |
|---|---|---|
| **Feature learning and selection** | | |
| Feature learning | N-gram | n=1, 2, 3, 5, and 7 |
| Feature selection | Significance level | Filtering alpha at $10^{-10}$, $10^{-8}$, $10^{-6}$, and $10^{-4}$ |
| **Classifiers** | | |
| ZeroR (Majority class predictor) | | (Control) |
| OneR algorithm | | |
| Naive Bayes | | |
| Logistic ridge regression | | Ridge estimator $\lambda=10^{-8}$ |
| Support vector machines | Both polynomial Kernel Radial basis function kernels | Linear with normalised attributes. $\varepsilon=10^{-12}$. C=1. Fitting logistic models for parameter estimation |
| Alternating decision trees | Number of boosting iteration | Expanding all search paths. 2, 4, 6, 8, 10, 12, 14, 16, 18, 20 |
| Random forests | Number of bagged trees | 5, 10, 15, 20, 25, 30, 35, 40, 45, 50 |
| **Meta-classifiers** | | |
| Bootstrap aggregation | Iteration | 10 |
| Model selection | | Selection by best classifier |

**Supplementary Figure S10 – Overall survival of patients in the discovery and validation cohorts by cancer type.**

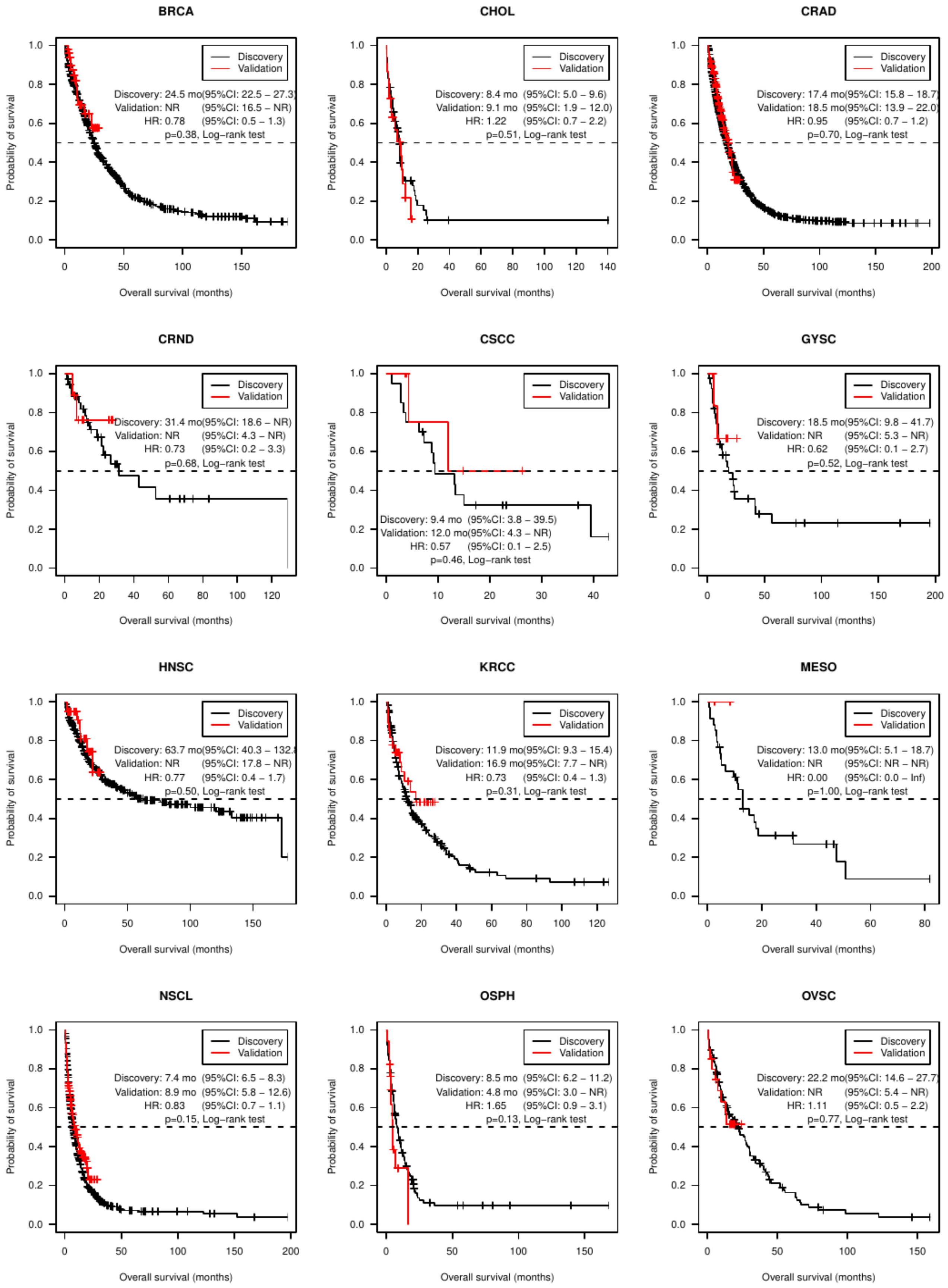

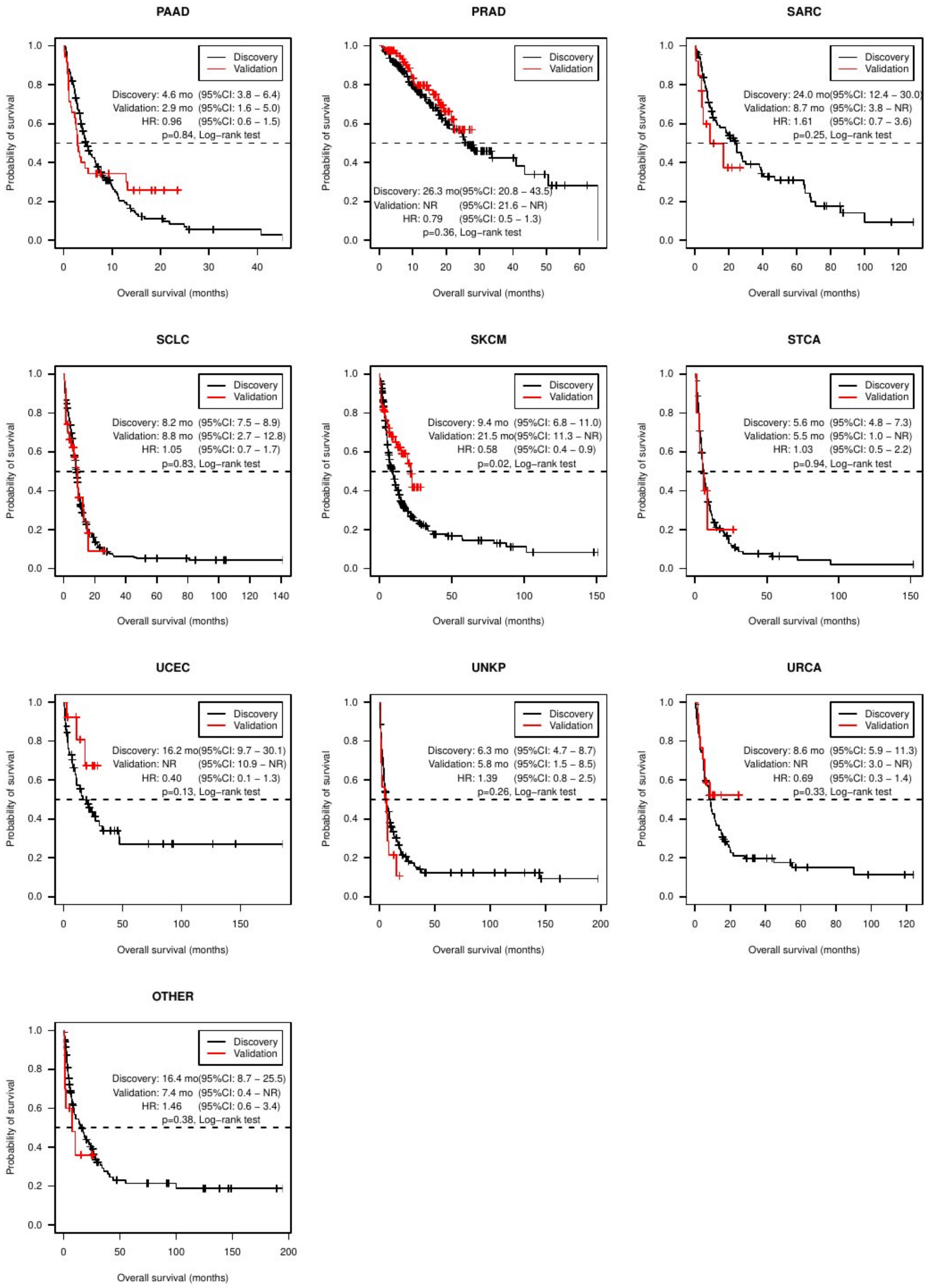
PAAD
Discovery
Validation
Discovery: 4.6 mo (95%CI: 3.8 – 6.4)
Validation: 2.9 mo (95%CI: 1.6 – 5.0)
HR: 0.96 (95%CI: 0.6 – 1.5)
p=0.84, Log−rank test
PRAD
Discovery
Validation
Discovery: 26.3 mo(95%CI: 20.8 – 43.5)
Validation: NR (95%CI: 21.6 – NR)
HR: 0.79 (95%CI: 0.5 – 1.3)
p=0.36, Log−rank test
SARC
Discovery
Validation
Discovery: 24.0 mo(95%CI: 12.4 – 30.0)
Validation: 8.7 mo (95%CI: 3.8 – NR)
HR: 1.61 (95%CI: 0.7 – 3.6)
p=0.25, Log−rank test
SCLC
Discovery
Validation
Discovery: 8.2 mo (95%CI: 7.5 – 8.9)
Validation: 8.8 mo (95%CI: 2.7 – 12.8)
HR: 1.05 (95%CI: 0.7 – 1.7)
p=0.83, Log−rank test
SKCM
Discovery
Validation
Discovery: 9.4 mo (95%CI: 6.8 – 11.0)
Validation: 21.5 mo(95%CI: 11.3 – NR)
HR: 0.58 (95%CI: 0.4 – 0.9)
p=0.02, Log−rank test
STCA
Discovery
Validation
Discovery: 5.6 mo (95%CI: 4.8 – 7.3)
Validation: 5.5 mo (95%CI: 1.0 – NR)
HR: 1.03 (95%CI: 0.5 – 2.2)
p=0.94, Log−rank test
UCEC
Discovery
Validation
Discovery: 16.2 mo(95%CI: 9.7 – 30.1)
Validation: NR (95%CI: 10.9 – NR)
HR: 0.40 (95%CI: 0.1 – 1.3)
p=0.13, Log−rank test
UNKP
Discovery
Validation
Discovery: 6.3 mo (95%CI: 4.7 – 8.7)
Validation: 5.8 mo (95%CI: 1.5 – 8.5)
HR: 1.39 (95%CI: 0.8 – 2.5)
p=0.26, Log−rank test
URCA
Discovery
Validation
Discovery: 8.6 mo (95%CI: 5.9 – 11.3)
Validation: NR (95%CI: 3.0 – NR)
HR: 0.69 (95%CI: 0.3 – 1.4)
p=0.33, Log−rank test
OTHER
Discovery
Validation
Discovery: 16.4 mo(95%CI: 8.7 – 25.5)
Validation: 7.4 mo (95%CI: 0.4 – NR)
HR: 1.46 (95%CI: 0.6 – 3.4)
p=0.38, Log−rank test
Probability of survival
Overall survival (months)

Comparison of overall survival in discovery and validation cohort in 22 cancer types. The survival of all cancer types were comparable across cancer types with the exception of cutaneous melanoma (9.4 v 21.5 months, HR 0.58, p=0.02), associated with public funding of pembrolizumab for treatment of metastatic cutaneous melanoma in September 2016.

**Supplementary Table S11 – Comparison of prognostic accuracy of ECOG performance status versus clinical text in predicting short to mid-term mortality in external validation cohort.**

| Div. | Cases | | | ECOG PS | | Text | | z | P | Sig [a] |
|---|---|---|---|---|---|---|---|---|---|---|
| | N | Died | Alive | AUC | SE | AUC | SE | | | |
| 14 | 441 | 7 | 434 | 0.876 | 0.085 | 0.862 | 0.089 | -0.16 | 0.87 | |
| 28 | 441 | 25 | 416 | 0.840 | 0.050 | 0.769 | 0.056 | -1.35 | 0.18 | |
| 42 | 441 | 49 | 392 | 0.780 | 0.040 | 0.820 | 0.038 | 1.03 | 0.30 | |
| 56 | 441 | 64 | 377 | 0.789 | 0.035 | 0.834 | 0.032 | 1.36 | 0.17 | |
| 70 | 441 | 74 | 367 | 0.783 | 0.033 | 0.829 | 0.030 | 1.44 | 0.15 | |
| 84 | 431 | 85 | 346 | 0.780 | 0.031 | 0.830 | 0.029 | 1.69 | 0.092 | |
| 112 | 414 | 102 | 312 | 0.758 | 0.030 | 0.816 | 0.027 | 2.02 | 0.043 | * |
| 140 | 404 | 110 | 294 | 0.744 | 0.030 | 0.822 | 0.026 | 2.79 | 0.0053 | * |
| 182 | 393 | 125 | 268 | 0.746 | 0.028 | 0.815 | 0.025 | 2.61 | 0.0091 | * |
| 273 | 348 | 159 | 189 | 0.738 | 0.027 | 0.794 | 0.024 | 2.26 | 0.024 | * |
| 364 | 328 | 178 | 150 | 0.740 | 0.027 | 0.783 | 0.025 | 1.66 | 0.096 | |

Note: Div: case dichotomisation interval (days). AUC: area under the ROC curve. SE: standard error estimated as by Hanley-McNiell method. ECOG PS: Eastern Cooperative Operative Group Performance Status. Mean difference in AUC across all divisions: 0.0367 ($\chi^2$=27.8, p=0.0019, Chi-square test, df=10). (a) Statistically significant by pairwise testing (*z*-test) at α=0.05.

**Supplementary Table S12 – Comparison of prognostic accuracy of ECOG performance status with and without clinical text in predicting short to mid-term mortality in patients with advanced cancers in the external validation cohort.**

| Div. | Cases | | | ECOG PS | | ECOG PS + Text | | z | P | Sig [a] |
|---|---|---|---|---|---|---|---|---|---|---|
| | N | Died | Alive | AUC | SE | AUC | SE | | | |
| 14 | 441 | 7 | 434 | 0.876 | 0.085 | 0.894 | 0.080 | 0.22 | 0.83 | |
| 28 | 441 | 25 | 416 | 0.840 | 0.050 | 0.810 | 0.053 | -0.58 | 0.56 | |
| 42 | 441 | 49 | 392 | 0.780 | 0.040 | 0.827 | 0.037 | 1.20 | 0.23 | |
| 56 | 441 | 64 | 377 | 0.789 | 0.035 | 0.833 | 0.032 | 1.33 | 0.18 | |
| 70 | 441 | 74 | 367 | 0.783 | 0.033 | 0.831 | 0.030 | 1.53 | 0.13 | |
| 84 | 431 | 85 | 346 | 0.780 | 0.031 | 0.836 | 0.028 | 1.88 | 0.060 | |
| 112 | 414 | 102 | 312 | 0.758 | 0.030 | 0.818 | 0.027 | 2.08 | 0.037 | * |
| 140 | 404 | 110 | 294 | 0.744 | 0.030 | 0.837 | 0.025 | 3.38 | 0.00073 | * |
| 182 | 393 | 125 | 268 | 0.746 | 0.028 | 0.825 | 0.025 | 3.00 | 0.0027 | * |
| 273 | 348 | 159 | 189 | 0.738 | 0.027 | 0.801 | 0.024 | 2.54 | 0.011 | * |
| 364 | 328 | 178 | 150 | 0.740 | 0.027 | 0.804 | 0.024 | 2.50 | 0.012 | * |

Note: Div: case dichotomisation interval. AUC: area under ROC curve. SE: standard error estimated as by Hanley-McNiell method. ECOG PS: Eastern Cooperative Operative Group Performance Status. Mean difference in AUC across all division: 0.0495 ($\chi^2$=31.34, p=0.00051, chi-square test with df=10). (a) Statistically significant by pairwise testing (*z*-test) at α=0.05.

**Supplementary Text S13 – Patient with multiple diagnoses of advanced cancers in the validation cohort**

In the validation cohort, two patients had more than one type of Stage IV cancer recorded in the registry: (1) a 59-year-old male with a metachronous diagnosis of squamous cell carcinoma of larynx and non-small cell lung carcinoma 12 months later. (2) A 68-year-old man with synchronous metastatic melanoma and rectal carcinoma.

**Supplementary Text S14 – Additional discussions**

We have conducted a comprehensive text mining analysis of clinical narratives from a tertiary oncology centre to systematically extract prognostic factors in advanced cancer patients. Conceptually, our computational approach captures the implicit expertise of oncologists that correlates to important prognostic landmarks across cancer types. Notably, the specific language with prognostic significance used by oncologists can be readily identified, and some previously unrecognised factors can lead to formulation of new hypotheses to inform future studies. We have also highlighted the application of the Word2Vec model to automatically group segments of free-text into meaningful clusters, allowing high-level interpretation, where words within a group can be potentially incorporated to refine prognostication assessment in patients with advanced cancer.

Combining machine learning models, we have shown that adding clinical narratives for prediction can improve accuracy of prognostic assessment than using ECOG performance status alone. Our findings reveal a unique opportunity to integrate EMR analysis to automate the prognostic tasks. The utility of a prognostic model from utilising clinical free-text at first consultation has value to help subsequent automated therapy planning. We showed that physician-authored text, when coupled with machine learning, can significantly improve prognostic accuracy in this patients group, compared to scoring methods purely based on performance status. The currently underutilized resource in both research and practice can thus be built to build automated predictive tools for improving quality of care. One potential application is to improve the stratification of patients for consideration of clinical trial participation referral, where accurate estimation of survival is required.

We propose that the machine learning approach presented here is highly adaptable to other clinical tasks. Conceptually, our results demonstrate how clinician's assessments can be integrated into computational models for predictions. While clinical narratives are often specific to sites and local practice, we have demonstrated that it is feasible to derive a "locally-built" model from EMR data, as validated in our cohort temporal validation. An interesting question remains unresolved, however, whether a predictive model derived using local data can be generalisable to another site of care (e.g., cancer centre). Furthermore, while population-based summary scoring methods may be easily used (e.g., ECOG score), the merit of adding in a data-driven approaches (e.g., based on clinical free-text and/or other clinicopathologic variables) was shown in our data where a combined approach has led to improved prognostic or predictive performance. How to integrate existing knowledge (e.g., score-based approach) and a data-driven approach is not well--examined to date.

Our data, however, has shed light on combining both models is feasible, suggesting the adding of raw patient-level data may offer an advantage over scoring rules derived using traditional epidemiology methods.

One intriguing observation was made: our informatics pipeline revealed predominantly clinical manifestations of cancer (e.g., symptoms and performance status) in the index analysis. The set of features identified strongly reflects the expertise of oncologists, where common disease-specific factors (e.g., histopathology features such as tumour grades, proliferation index, or receptor status in invasive carcinoma of breast) were less significant (Supplementary Dataset S3). While this is largely due to the application of statistical adjustment to account for heterogeneity of cancer types (i.e., normalised OS), the true difference is likely determined by to the types of corpus (i.e., clinic letters), such that further examination of pathology or radiology reports may yield a different set of prognostic factors that may otherwise not be revealed by clinical correspondence alone.

Our studies have several notable strengths. First, the clinical prognostic factors extracted by TEPAPA are explicit and readily interpretable, and its identification can rapidly provide insights into a clinical dataset. Our method also enables rapid, unbiased screening of potential prognostic covariates from clinical free-text to accelerate the hypothesis generation step in formulating observational studies. Second, we have demonstrated how voluminous free-text in EMR can be readily repurposed to create an "end-to-end" predictive tool with potential clinical application. This opens up opportunities to create predictive tools for certain outcomes of interest where covariates may not be fully known. Third, while some "black box" predictive models, such as artificial neural networks, could provide better accuracy in prediction, the transparency of our informatics pipeline allows direct inspection of possible covariates that improves the interpretability of the model.

Several limitations are also noted where further research is required. First, the results presented here are limited to the first oncology consultation. While it guarantees within-subject statistical independence, construction of a temporal model is useful for identifying key factors to predict a patient's disease trajectory, hence providing risk stratification beyond the first clinic visit. Second, up to 40% of EMR were not available for analysis, as some clinic letters were not accessible from the departmental registry outside the index hospital, highlighting the real-world challenge of repurposing EMR free-text in practice. Sample size alone, however, did not preclude discovery of plausible prognostic factors or compromised classifier performance as demonstrated in our results. Third, more sophisticated natural language processing (NLP) methods are available in clinical document classification [S14.1]. Nonetheless, data-driven models with a "white box" feature extraction method allows explanation as we have demonstrated in our work. Fourth, the main objective of our work was to evaluate whether clinical text may enhance the prediction of in conjunction with stratification based on performance status; other clinical and demographic factors, such as physiological parameters, clinicopathologic variables, and age, were not assessed. It is plausible that the prognostic accuracy can be further extended by including other available variables from the EMR in this patient group; future work should also be carried out to evaluate its utility and impact on clinical practice. Fifth, one unresolved challenge common to all computational models, is on the fairness of algorithms with respect to marginalised groups. It is known that minority populations (e.g., ethnicity or rare cancer types) can be under-represented in the training data, such that they are at risk of being less accurately identified by data-driven modelling [S14.2]. Governance strategies have been recommended as a potential solution to mitigate this problem [S14.3]. We further propose that involvement of Indigenous researchers and others with expertise in health equity is crucial to mitigate the risk of biases.

***References***


[S14.1] Spasic I, Nenadic G. Clinical Text Data in Machine Learning: Systematic Review. JMIR Med Inform 2020; 8:e17984.

[S14.2] Blasimme A, Vayena E. The Ethics of AI in Biomedical research, patient care and public health. Patient Care and Public Health (April 9, 2019). Oxford Handbook of Ethics of Artificial Intelligence, Forthcoming. 2019 Apr 9.

[S14.3] Reddy S, Allan S, Coghlan S, Cooper P. A governance model for the application of AI in health care. J Am Med Inform Assoc 2020; 27:491-497.

**Supplementary Figure S15 – Principal component analysis of the 20-dimension vectors of the unigrams discovered in the index analysis.**

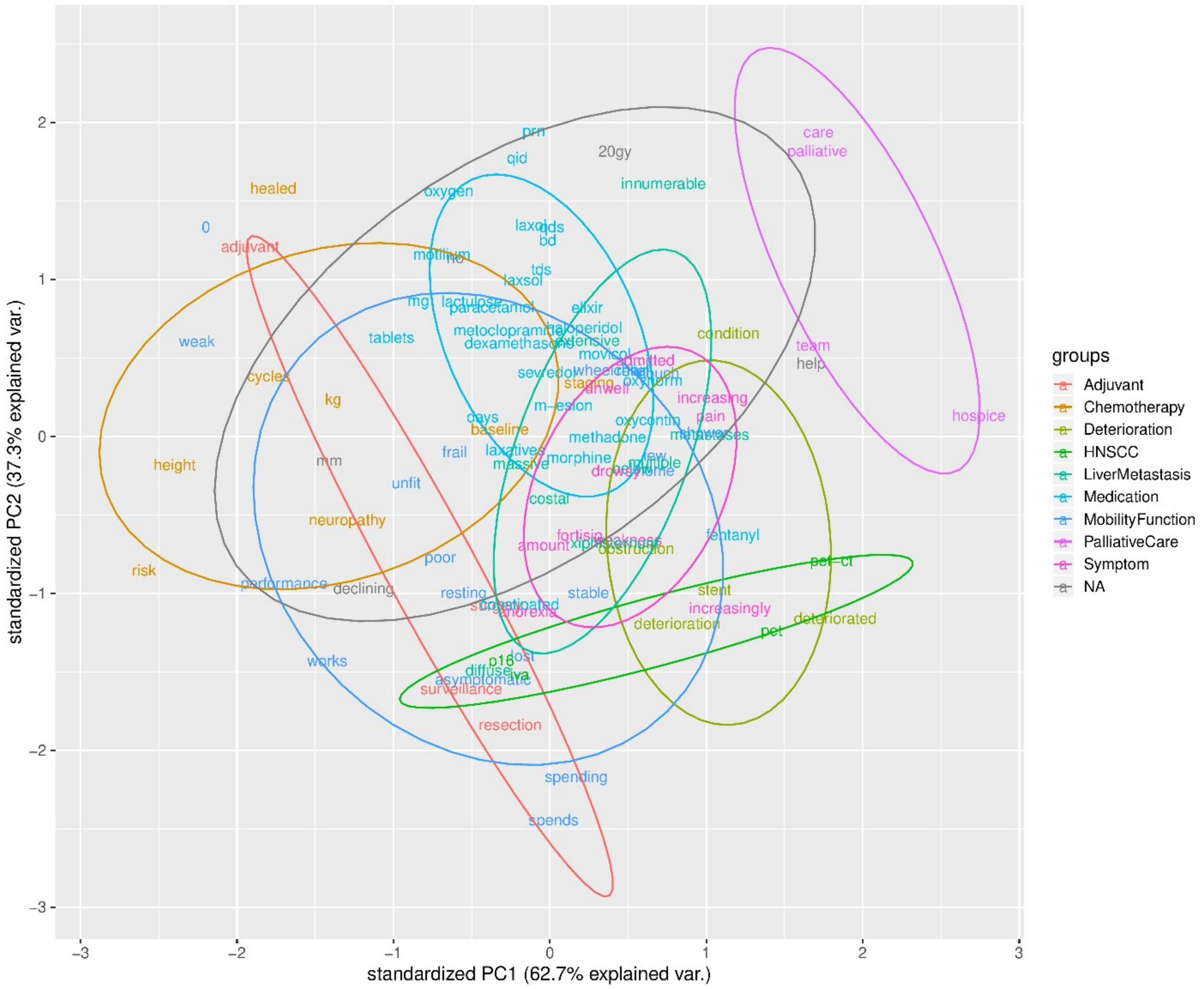


Principal component analysis of the words clusters of closely-related unigrams

**Supplementary Figure S16 – Overall survivals in Maori and non-Maori patients of entire cohort**

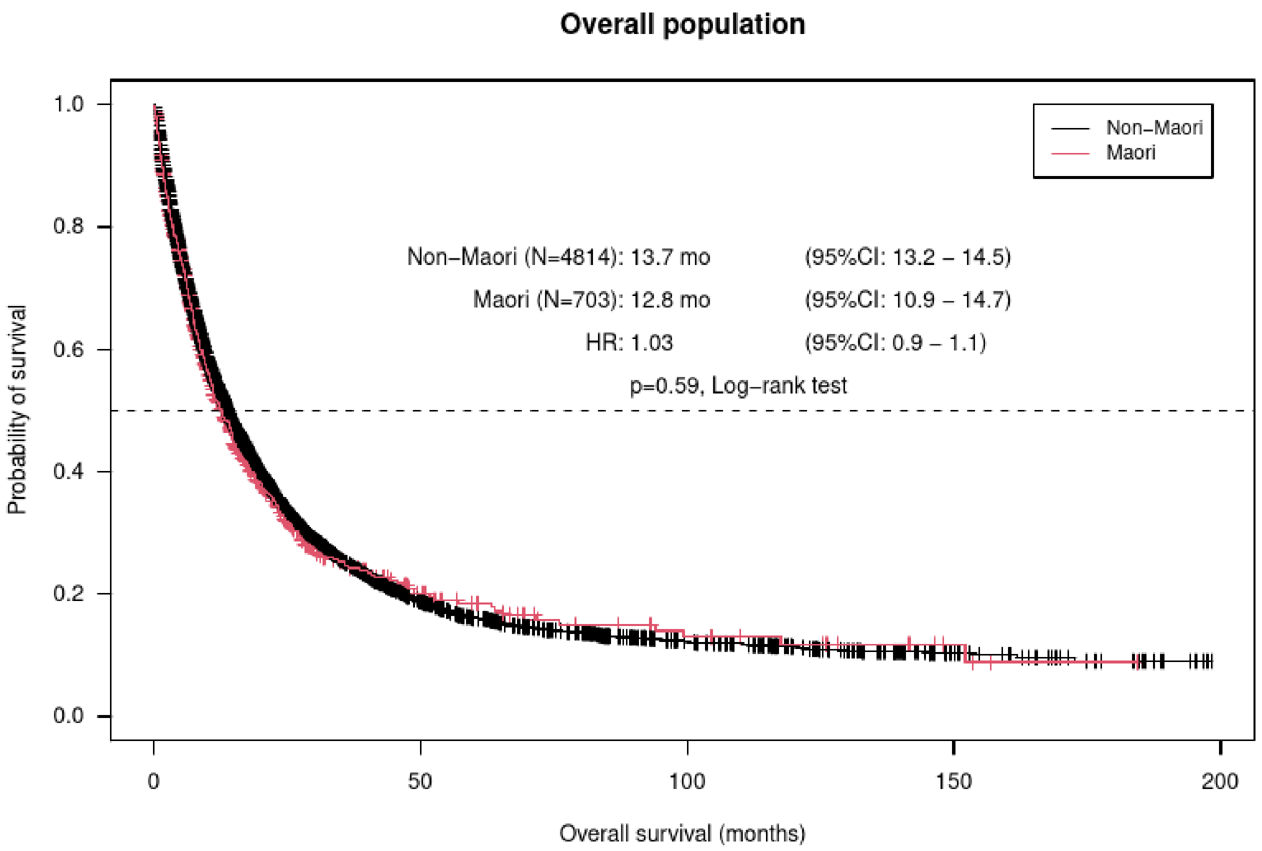


The Kaplan-Meier analyses showed overall survival of 12.8 months and 13.7 months in Maori vs Non-Maori patients respectively (HR: 1.03; 95% CI: 0.93-1.13). No statistically significant difference in durations of overall survival was seen with respect to ethnicity of patients (Maori v non-Maori) in the combined cohort (discovery + validation).

**Supplementary Figure S17 – Overall survivals in Maori and non-Maori patients of entire cohort by cancer types**

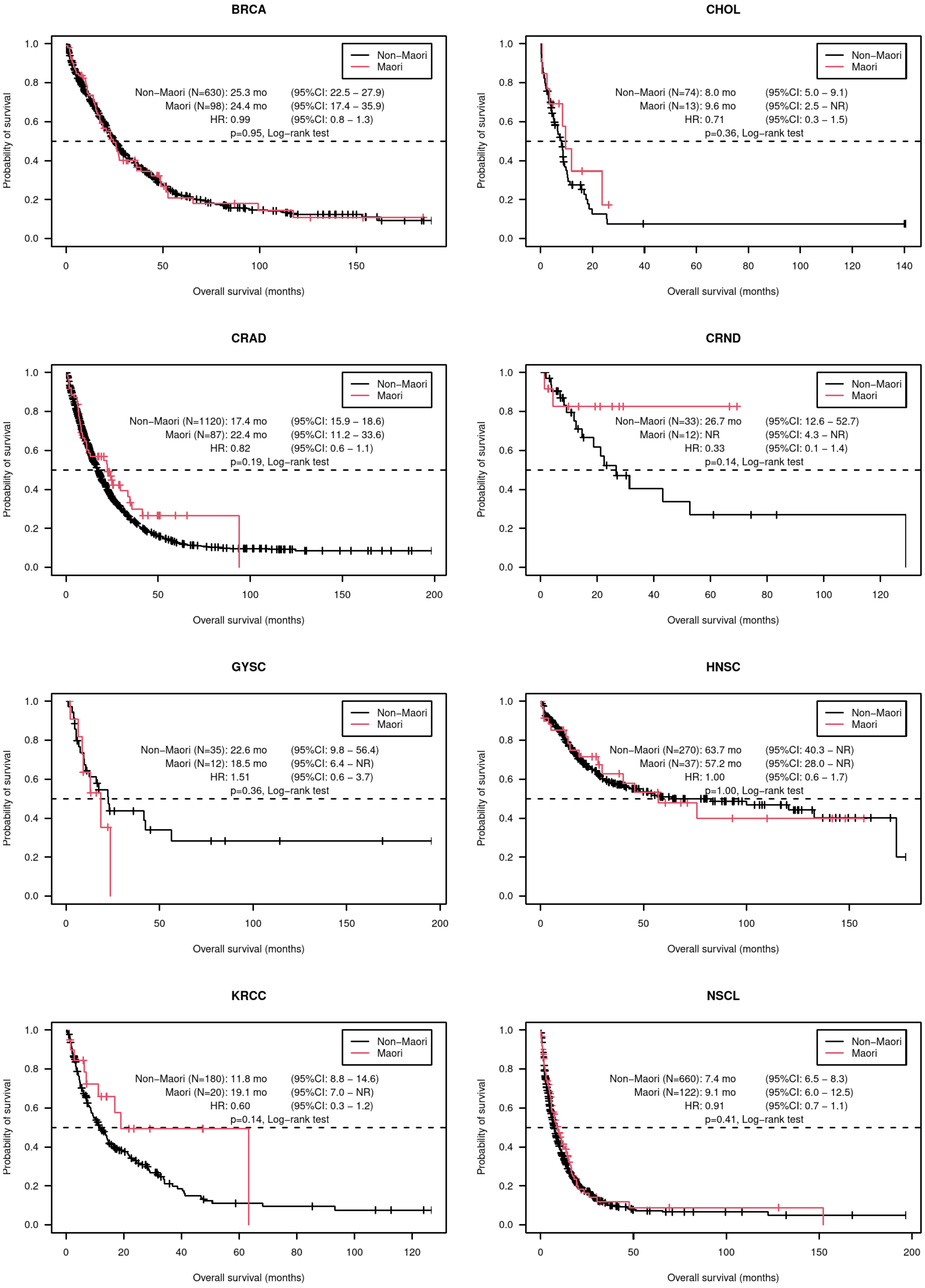

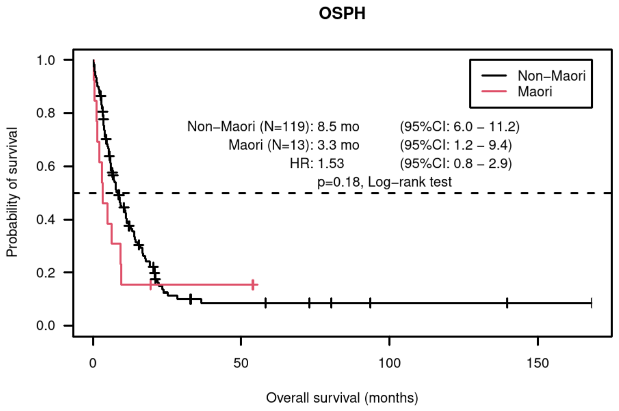
OSPH
Non−Maori
Maori
Non−Maori (N=119): 8.5 mo (95%CI: 6.0 − 11.2)
Maori (N=13): 3.3 mo (95%CI: 1.2 − 9.4)
HR: 1.53 (95%CI: 0.8 − 2.9)
p=0.18, Log−rank test
Probability of survival
Overall survival (months)
1.0
0.8
0.6
0.4
0.2
0.0
0
50
100
150

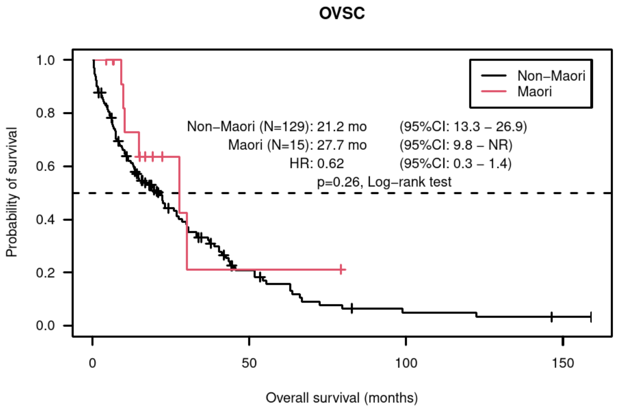
OVSC
Non−Maori
Maori
Non−Maori (N=129): 21.2 mo (95%CI: 13.3 − 26.9)
Maori (N=15): 27.7 mo (95%CI: 9.8 − NR)
HR: 0.62 (95%CI: 0.3 − 1.4)
p=0.26, Log−rank test
Probability of survival
Overall survival (months)
1.0
0.8
0.6
0.4
0.2
0.0
0
50
100
150

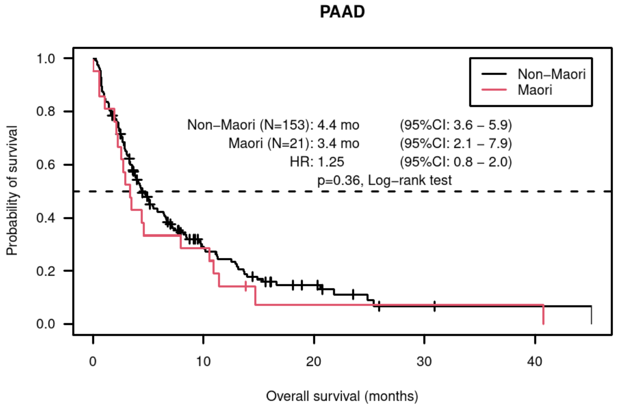
PAAD
Non−Maori
Maori
Non−Maori (N=153): 4.4 mo (95%CI: 3.6 − 5.9)
Maori (N=21): 3.4 mo (95%CI: 2.1 − 7.9)
HR: 1.25 (95%CI: 0.8 − 2.0)
p=0.36, Log−rank test
Probability of survival
Overall survival (months)
1.0
0.8
0.6
0.4
0.2
0.0
0
10
20
30
40

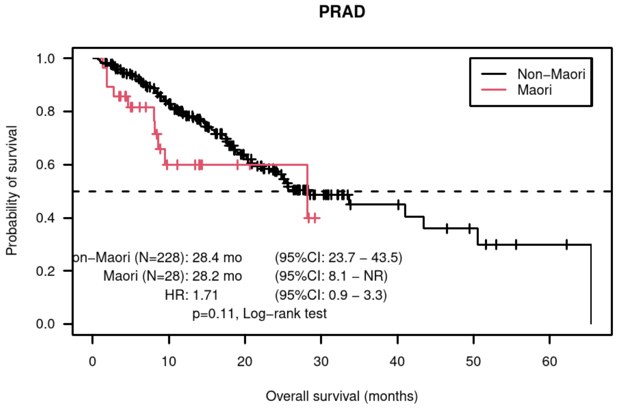
PRAD
Non−Maori
Maori
on−Maori (N=228): 28.4 mo (95%CI: 23.7 − 43.5)
Maori (N=28): 28.2 mo (95%CI: 8.1 − NR)
HR: 1.71 (95%CI: 0.9 − 3.3)
p=0.11, Log−rank test
Probability of survival
Overall survival (months)
1.0
0.8
0.6
0.4
0.2
0.0
0
10
20
30
40
50
60

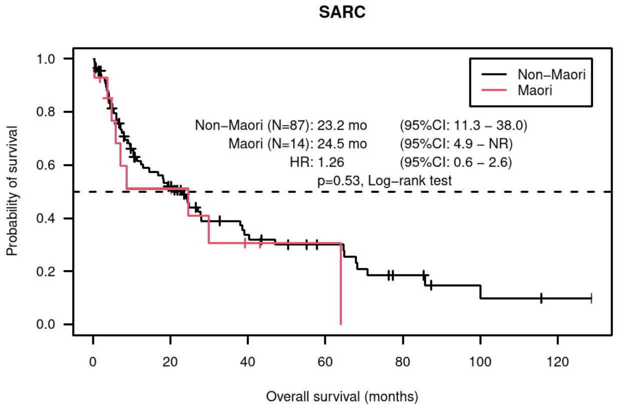
SARC
Non−Maori
Maori
Non−Maori (N=87): 23.2 mo (95%CI: 11.3 − 38.0)
Maori (N=14): 24.5 mo (95%CI: 4.9 − NR)
HR: 1.26 (95%CI: 0.6 − 2.6)
p=0.53, Log−rank test
Probability of survival
Overall survival (months)
1.0
0.8
0.6
0.4
0.2
0.0
0
20
40
60
80
100
120

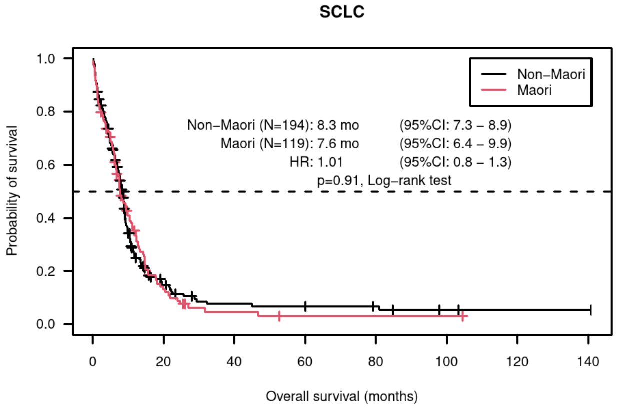
SCLC
Non−Maori
Maori
Non−Maori (N=194): 8.3 mo (95%CI: 7.3 − 8.9)
Maori (N=119): 7.6 mo (95%CI: 6.4 − 9.9)
HR: 1.01 (95%CI: 0.8 − 1.3)
p=0.91, Log−rank test
Probability of survival
Overall survival (months)
1.0
0.8
0.6
0.4
0.2
0.0
0
20
40
60
80
100
120
140

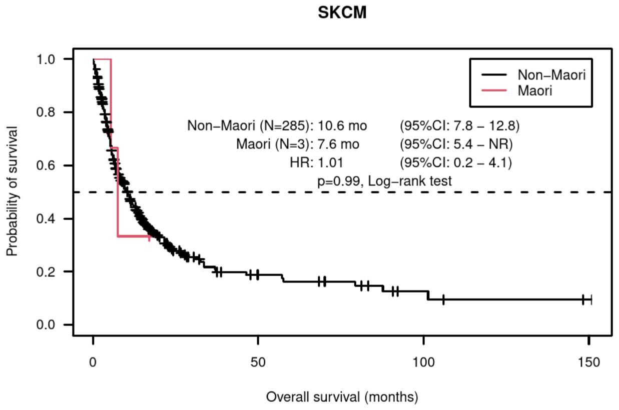
SKCM
Non−Maori
Maori
Non−Maori (N=285): 10.6 mo (95%CI: 7.8 − 12.8)
Maori (N=3): 7.6 mo (95%CI: 5.4 − NR)
HR: 1.01 (95%CI: 0.2 − 4.1)
p=0.99, Log−rank test
Probability of survival
Overall survival (months)
1.0
0.8
0.6
0.4
0.2
0.0
0
50
100
150

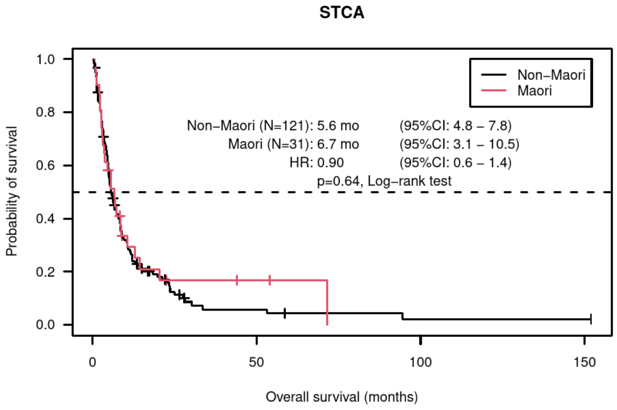
STCA
Non−Maori
Maori
Non−Maori (N=121): 5.6 mo (95%CI: 4.8 − 7.8)
Maori (N=31): 6.7 mo (95%CI: 3.1 − 10.5)
HR: 0.90 (95%CI: 0.6 − 1.4)
p=0.64, Log−rank test
Probability of survival
Overall survival (months)
1.0
0.8
0.6
0.4
0.2
0.0
0
50
100
150

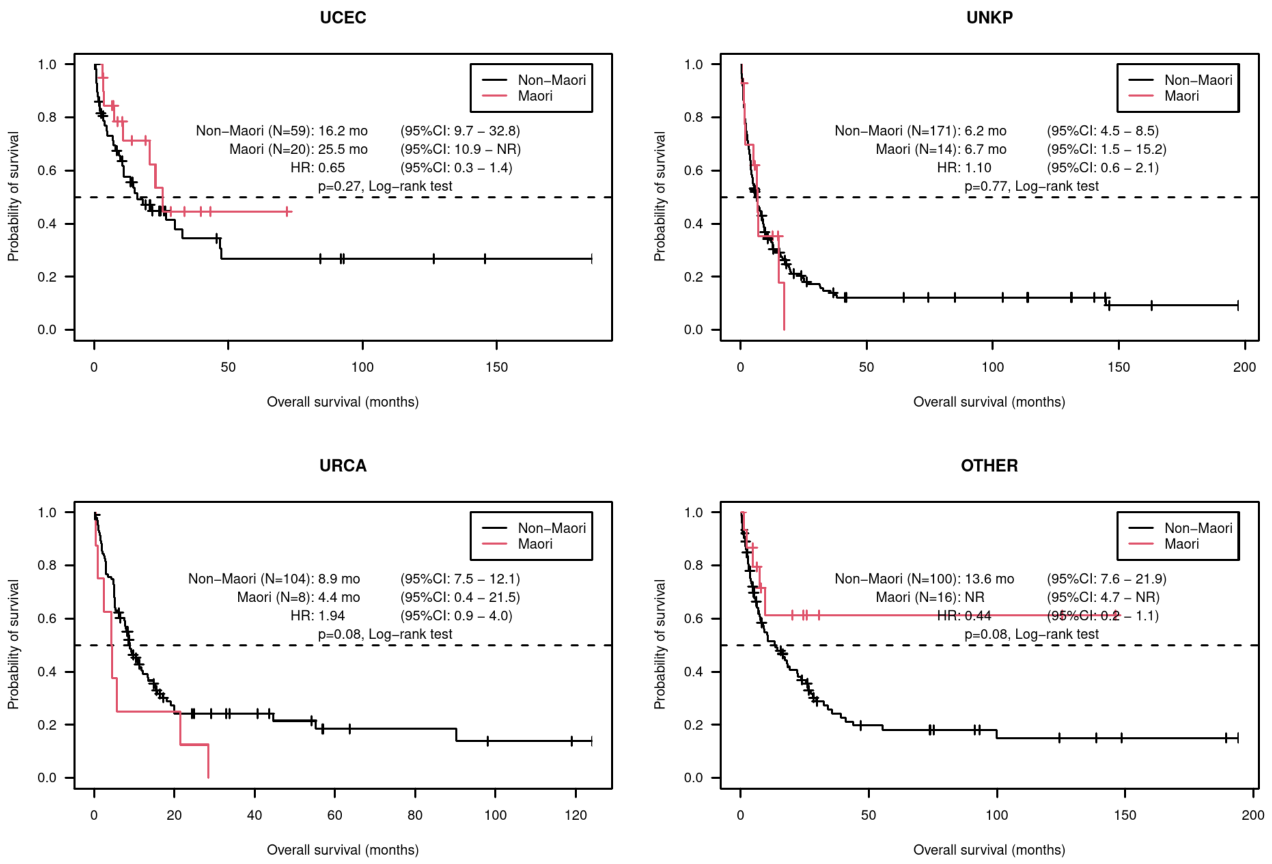


The Kaplan-Meier analyses showed no statistically significant difference in durations of overall survival by ethnicity of patients (Maori v non-Maori) in the combined cohort (discovery + validation) across twenty groups of cancer types. Abbreviations: BRCA: breast cancer; CHOL: biliary cancers; CRAD: colorectal cancer; CRND: neuroendocrine tumours; GYSC: cervical, vaginal, and vulvar cancers; HNSC: head and neck squamous cell carcinoma; KRCC: Kidney cancers; NSCL: Lung non-small cell carcinomas; OSPH: Oesophageal cancers; OVSC: Ovarian cancers; PAAD: Pancreatic cancers; PRAD: Prostate adenocarcinoma; SARC: Sarcomas; SCLC: Lung small-cell carcinoma; SKCM: Melanoma; STCA: Gastric cancer; UCEC: Uterine cancers; UNKP: cancer of unknown primary; URCA: Bladder cancers; Other: rare and other tumour types.

**Supplementary Table S18 – STrengthening the Reporting of OBservational studies in Epidemiology (STROBE) statement**

| | Item No | Recommendation | Page No. |
|---|---|---|---|
| **Title and abstract** | 1 | (*a*) Indicate the study's design with a commonly used term in the title or the abstract | Retrospective cohort study |
| | | (*b*) Provide in the abstract an informative and balanced summary of what was done and what was found | Text provided, in accordance to the journal's formatting |
| **Introduction** | | | |
| Background/rationale | 2 | Explain the scientific background and rationale for the investigation being reported | Paragraph 1 |
| Objectives | 3 | State specific objectives, including any prespecified hypotheses | Paragraph 2 |
| **Methods** | | | |
| Study design | 4 | Present key elements of study design early in the paper | Paragraph 2 |
| Setting | 5 | Describe the setting, locations, and relevant dates, including periods of recruitment, exposure, follow-up, and data collection | Paragraph 3 |
| Participants | 6 | (*a*) Give the eligibility criteria, and the sources and methods of selection of participants. Describe methods of follow-up | Paragraph 3 |
| | | (*b*) For matched studies, give matching criteria and number of exposed and unexposed | Not applicable |
| Variables | 7 | Clearly define all outcomes, exposures, predictors, potential confounders, and effect modifiers. Give diagnostic criteria, if applicable | Paragraph 4<br>Confounder adjustment by stratification (nOS). |
| Data sources/ measurement | 8* | For each variable of interest, give sources of data and details of methods of assessment (measurement). Describe comparability of assessment methods if there is more than one group | Paragraph 4 |
| Bias | 9 | Describe any efforts to address potential sources of bias | Paragraph 5 |
| Study size | 10 | Explain how the study size was arrived at | Retrospective study, entire cohort included. |
| Quantitative variables | 11 | Explain how quantitative variables were handled in the analyses. If applicable, describe which groupings were chosen and why | Not applicable. |
| Statistical methods | 12 | (*a*) Describe all statistical methods, including those used to control for confounding | Online methods |
| | | (*b*) Describe any methods used to examine subgroups and interactions | Paragraph 5 |
| | | (*c*) Explain how missing data were addressed | Online methods |
| | | (*d*) If applicable, explain how loss to follow-up was addressed | Online methods |
| | | (*e*) Describe any sensitivity analyses | 11 time points external validation. |
| **Results** | | | |
| Participants | 13* | (a) Report numbers of individuals at each stage of study—eg numbers potentially eligible, examined for eligibility, confirmed eligible, included in the study, completing follow-up, and analysed | Figure 1. |
| | | (b) Give reasons for non-participation at each stage | Figure 1. |

| | | | |
|---|---|---|---|
| | | (c) Consider use of a flow diagram | Figure 1. |
| Descriptive data | 14* | (a) Give characteristics of study participants (eg demographic, clinical, social) and information on exposures and potential confounders | Table 1. |
| | | (b) Indicate number of participants with missing data for each variable of interest | Figure 1. |
| | | (c) Summarise follow-up time (eg, average and total amount) | Median follow-up time provided. |
| Outcome data | 15* | Report numbers of outcome events or summary measures over time | Table 1. |
| Main results | 16 | (*a*) Give unadjusted estimates and, if applicable, confounder-adjusted estimates and their precision (eg, 95% confidence interval). Make clear which confounders were adjusted for and why they were included | Uncertainties provided. |
| | | (*b*) Report category boundaries when continuous variables were categorized | Not applicable. |
| | | (*c*) If relevant, consider translating estimates of relative risk into absolute risk for a meaningful time period | Not applicable. |
| Other analyses | 17 | Report other analyses done—eg analyses of subgroups and interactions, and sensitivity analyses | Orthogonal analyses provided. |
| **Discussion** | | | |
| Key results | 18 | Summarise key results with reference to study objectives | |
| Limitations | 19 | Discuss limitations of the study, taking into account sources of potential bias or imprecision. Discuss both direction and magnitude of any potential bias | Supplementary text. |
| Interpretation | 20 | Give a cautious overall interpretation of results considering objectives, limitations, multiplicity of analyses, results from similar studies, and other relevant evidence | Paragraph 9. Supplementary text. |
| Generalisability | 21 | Discuss the generalisability (external validity) of the study results | Paragraph 10. |
| **Other information** | | | |
| Funding | 22 | Give the source of funding and the role of the funders for the present study and, if applicable, for the original study on which the present article is based | Provided. |

**Supplementary Table S19 – REporting recommendations for tumour MARKer prognostic studies (REMARK) statement**

| | Item to be reported | Page no. |
|---|---|---|
| **INTRODUCTION** | | |
| 1 | State the marker examined, the study objectives, and any pre-specified hypotheses. | Paragraph 2 |
| **MATERIALS AND METHODS** | | |
| *Patients* | | |
| 2 | Describe the characteristics (e.g., disease stage or co-morbidities) of the study patients, including their source and inclusion and exclusion criteria. | Paragraph 3. Figure 1. |
| 3 | Describe treatments received and how chosen (e.g., randomized or rule-based). | Not applicable |
| *Specimen characteristics* | | |
| 4 | Describe type of biological material used (including control samples) and methods of preservation and storage. | Not applicable |
| *Assay methods* | | |
| 5 | Specify the assay method used and provide (or reference) a detailed protocol, including specific reagents or kits used, quality control procedures, reproducibility assessments, quantitation methods, and scoring and reporting protocols. Specify whether and how assays were performed blinded to the study endpoint. | Online methods |
| *Study design* | | |
| 6 | State the method of case selection, including whether prospective or retrospective and whether stratification or matching (e.g., by stage of disease or age) was used. Specify the time period from which cases were taken, the end of the follow-up period, and the median follow-up time. | Paragraph 3 |
| 7 | Precisely define all clinical endpoints examined. | Paragraph 3 |
| 8 | List all candidate variables initially examined or considered for inclusion in models. | Paragraph 4 & Supplementary material |
| 9 | Give rationale for sample size; if the study was designed to detect a specified effect size, give the target power and effect size. | Not applicable |
| *Statistical analysis methods* | | |
| 10 | Specify all statistical methods, including details of any variable selection procedures and other model-building issues, how model assumptions were verified, and how missing data were handled. | Online methods |
| 11 | Clarify how marker values were handled in the analyses; if relevant, describe methods used for cutpoint determination. | Online methods |
| **RESULTS** | | |
| *Data* | | |
| 12 | Describe the flow of patients through the study, including the number of patients included in each stage of the analysis (a diagram may be helpful) and reasons for dropout. Specifically, both overall and for each subgroup extensively examined report the numbers of patients and the number of events. | Figure 1 |
| 13 | Report distributions of basic demographic characteristics (at least age and sex), standard (disease-specific) prognostic variables, and tumor marker, including numbers of missing values. | Table 1. |
| *Analysis and presentation* | | |
| 14 | Show the relation of the marker to standard prognostic variables. | Figure 2. |
| 15 | Present univariable analyses showing the relation between the marker and outcome, with the estimated effect (e.g., hazard ratio and survival probability). Preferably provide similar analyses for all other variables being analyzed. For the effect of a tumor marker on a time-to-event outcome, a Kaplan-Meier plot is recommended. | Figure 2. Supplementary Table |
| 16 | For key multivariable analyses, report estimated effects (e.g., hazard ratio) with confidence intervals for the marker and, at least for the final model, all other variables in the model. | Provided |
| 17 | Among reported results, provide estimated effects with confidence intervals from an analysis in which the marker and standard prognostic variables are included, regardless of their statistical significance. | Provided (comparison with ECOG PS) |

| | | |
|---|---|---|
| 18 | If done, report results of further investigations, such as checking assumptions, sensitivity analyses, and internal validation. | 11 time points + external validation. |
| **DISCUSSION** | | |
| 19 | Interpret the results in the context of the pre-specified hypotheses and other relevant studies; include a discussion of limitations of the study. | Supplementary Text |
| 20 | Discuss implications for future research and clinical value. | Paragraph 10; Supplementary text |
